# Changes in the physical behaviour of older adults during the 13 weeks GOTO intervention explain a boost in immuno-metabolic health

**DOI:** 10.1101/2023.11.26.23299026

**Authors:** Stylianos Paraschiakos, Fatih A. Bogaards, Arno Knobbe, P. Eline Slagboom, Marian Beekman

## Abstract

**Background:** The aging population faces numerous health challenges, with sedentary behavior and decreased physical activity being paramount. We explore the physical behaviour of older adults in the GOTO combined lifestyle intervention study and its related immuno-metabolic health effects.

**Methods:** The research utilized accelerometers and machine learning to assess physical activity behaviours during a 13-week program of increased physical activity and decreased calorie intake. Subsequently, the association of variation in physical behaviour with immuno-metabolic health parameters is investigated cross-sectionally at baseline and longitudinally using sex-stratified linear regression and linear mixed regression respectively.

**Results:** Participants exhibited physical behaviors similar to their age-matched peers from the UK-Biobank. Interestingly, gender-based differences were evident, with men and women showing distinct daily physical behavioural patterns. At baseline, a positive correlation was found between higher physical behavior and a healthier immune-metabolic profile, particularly in men. The longitudinal changes depict an overall boost in activity levels, predominantly among women. While increasing general activity and engaging in intense exercises proved advantageous for physical health, the immune-metabolic health benefits were more pronounced in men.

**Conclusion:** The short-term GOTO intervention underscores the significance of regular physical activity in promoting healthy aging even in middle to older age. Gender differences in behavior and health benefits deserve much more attention though. Our results advocate the broader implementation of such programs and emphasize the utility of technology, like accelerometers and machine learning, in both monitoring and promoting active lifestyles among older adults.

## 1 Introduction

With overall increased life expectancy, the number of older adults is growing, with implications for many sectors of society, including financial and labour markets as well as health care systems [1, 2, 3]. Unhealthy lifestyle choices increase the risk of age-related diseases and multi-morbidity, but these risks can be mitigated through lifestyle and behavioural changes [4, 5, 6]. To stimulate healthy ageing, it is important to monitor both risk-increasing and protective factors [7, 8]. One such protective factor is active physical behaviour, which has been shown to lower the risk of diabetes, cardiovascular disease, cancer, and multi-morbidity, to name but a few [9]. Thus, monitoring physical behaviour as a protective factor in older adults is of high importance in the healthy ageing process.

Physical behaviour (PB) refers to the various physical activities (PA) that an individual engages in, as measured over an extended period of weeks to months. Longitudinal [10, 11, 12] and cross-sectional [13, 14, 15] studies have linked different PAs to healthy ageing. These studies, until recently, have relied on self-reported questionnaires [16], which have been found to have reporting biases [17, 18]. Nowadays, studies increasingly employ wearable technologies such as accelerometers to monitor PA objectively. One of the most common techniques in PA research is the estimation of low, medium, and vigorous PA levels based on acceleration magnitude cut-offs. Such PA levels can then be quantified into scores, and time spent on sedentary PA (SPA), light PA (LPA), or moderate to vigorous PA (MVPA). As a result, these scores can be associated with health and ageing markers. In general, studies in older adults have found that both short and prolonged MVPA episodes throughout the day associate with healthier body composition (lower weight or fat percentages) [19], can improve cardio-metabolic health [9, 20, 21, 22], and lower the risk of multi-morbidity [23].

Classifying acceleration magnitude in low, medium, or vigorous PA levels using cut-offs might be a straightforward step, however, these cut-offs are population-dependent [24] and do not take into account valuable information such as the fluctuations and patterns of the accelerometer signal. Furthermore, these categories can be challenging to translate into clear public health recommendations for older adults [13], who are underrepresented in PA research [25]. Behavioural information like the time spent performing specific activities, e.g. sitting, standing, walking, or cycling, would make PA studies better translatable into guidelines for physical behaviour (PB). To extract such information from accelerometer signals and gain a more accurate understanding of the nature and health benefits of PA behaviours, particularly among older adults, artificial intelligence methods such as human activity recognition can be applied. These methods use machine learning algorithms to classify specific patterns in the accelerometer signal into different types of activities [26]. While these models have been successfully used in other studies [13], they have not yet been applied in the context of health benefits for older adults.

Given the heterogeneity in the physiology of older adults, it is currently not known how long and at what intensity older adults of varying health statuses, sex, and age need to increase their daily PA to notice health benefits. To address this knowledge gap, one should follow older adults during a baseline PA period and a period of increased PA while simultaneously assessing any health changes. PA monitoring can be achieved by using wearable technology and the interpretation of PA-related behaviour by applying AI algorithms [27]. Moreover, the ensuing health benefits can be monitored by observing the changes in health indicators predictive of disease risk, for instance, blood-based immune and metabolic health factors linked with virtually all diseases of old age [28]. Such studies can contribute to the development of tailored recommendations for effectively delaying the onset of disease and improving health in older adults.

To investigate the relationship between changes in PB and those in immune and metabolic health in older adults, we explored a subgroup of the Growing Old Together (GOTO) lifestyle intervention study [29]. GOTO was a 13-week lifestyle intervention study in which older participants with healthy to overweight body mass index aimed to increase physical activity energy expenditure by 12.5% while decreasing their energy intake by 12.5%. In the GOTO subgroup studied in detail in this paper, PB parameters were estimated by combining ankle and wrist acceleration magnitude and human activity recognition, using the LARA algorithm [26]. First, as a confirmatory step, we explore at baseline the PB of the participants, which time of the day they are most active, and whether that is similar for men and women. Second, we correlate the time spent on certain activities at certain hours of the day with a selected set of immune and metabolic health parameters at the baseline of the 13-week lifestyle intervention. Third, we determine the changes in PB due to the GOTO lifestyle intervention, and the effect of that on the available health parameters. Finally, we explore whether the response of the health parameters can be explained by the change in physical behaviour (PB) and whether the response differs per sex.

## 2 Methods

### 2.1 GOTO lifestyle intervention

The current study utilizes a subset from a larger study: the single-arm GOTO lifestyle intervention trial that was conducted between 2012 and 2014 [29]. The trial design for GOTO received approval from the medical ethical committee at Leiden University Medical Center (*P* 11.187) and all participants provided written informed consent. The completion of the study adhered to all applicable guidelines and regulations and was registered as *NTR*3499 at the Dutch Trial Register ^7^.

The GOTO study implemented a 13-week lifestyle program with the goal of achieving a 12.5% caloric restriction and 12.5% increase in energy expenditure through an increase in physical activity while participants were under the supervision of a dietitian and a physiotherapist. Figure 1 illustrates the study design. The study recruited 164 healthy older adults from the Netherlands, with an average age of 63 years and a BMI range of 22.3 − 35.2 kg/m^2^. None of the participants had diabetes or any other significant medical conditions that could impact their weight or body composition, including active cancers. Additionally, participants provided a report of their current medication use from their pharmacist.

**Figure 1:**
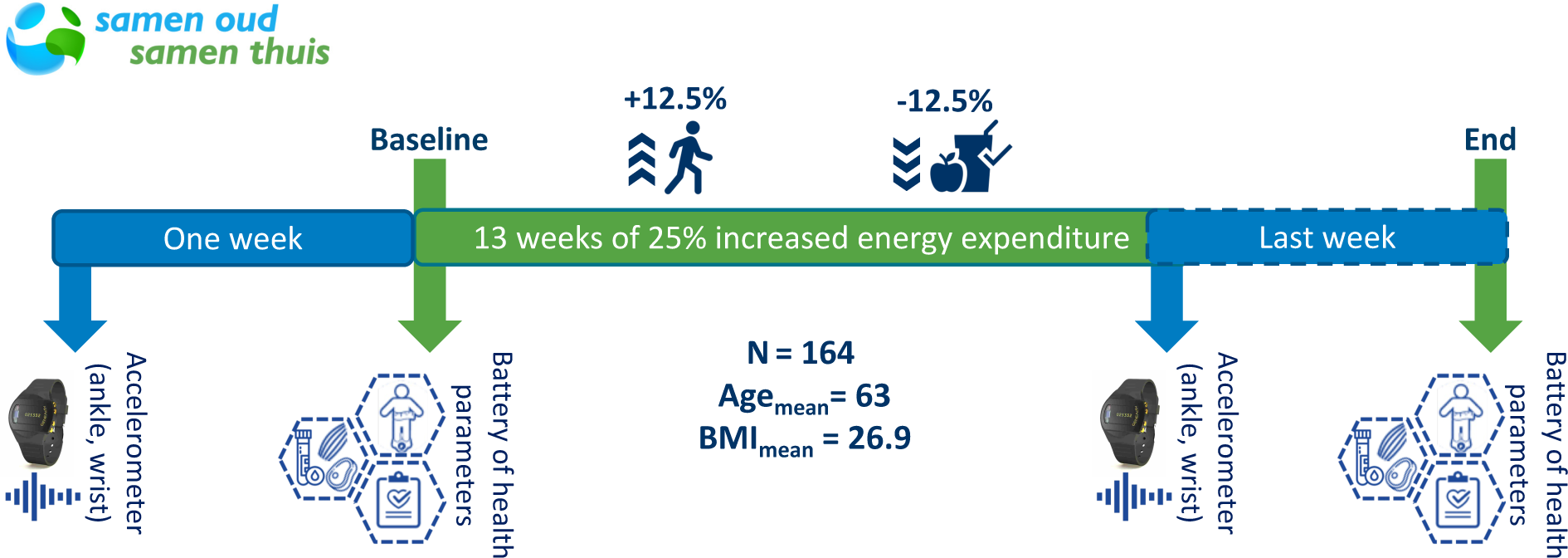
The Growing Old TOgether intervention study design.

#### 2.1.1 Collection of health parameters

The health parameters in the GOTO study, and thus our selected subgroup, were measured before and after the intervention and included anthropometric parameters of body composition (weight, height, and waist and hip circumference) [29] and DEXA scan features (body fat, lean mass, trunk fat, etc.) [30], blood pressure, inflammation and metabolism-related biochemical blood markers, and metabolomics features like ^1^H-NMR metabolomic (Nightingale Health) [29].

### 2.2 Physical behaviour measurements

Physical activity of the older adults was quantified using two GENEActiv^8^ triaxial accelerometers placed on the non-dominant wrist and ankle of each participant. This approach of using both upper and lower limb movement measurements provides a comprehensive picture of the overall physical activity patterns of the participants [26]. Participants were required to wear the devices for a minimum of one week (7 consecutive, 24-hour, days), both before the start and at the 13th week of the intervention (see Figure 1). Acceleration was measured at 85.7 Hz and it was reported in relation to gravity^9^. The Euclidean Norm Minus One (ENMO) [31] was then calculated to quantify movement per second from both devices, reported in milligravity (mg) per second with negative values rounded to zero. Following that, a quality control step took place where non-wearing periods were identified based on the ENMO variation per hour for both ankle and wrist.

To analyze the impact of the lifestyle intervention on the physical behaviour of the participants, besides ENMO we employed metrics from the LARA algorithm [26]. LARA is an AI model for human activity recognition that was developed and validated in older adults. LARA uses machine learning techniques to predict the specific activities that participants performed before and after the intervention, such as walking, household tasks, sitting, cycling, etc. The LARA algorithm produces activity predictions per second. The selected 15 different types of daily PA to predict are: lying down (left or right), sitting (on a chair, or sofa, with both legs on and parallel to the ground and standing still), household activities (dishwashing, stacking selves, vacuum cleaning), walking (slow, normal, fast, stairs up, stepping) and cycling. This approach allowed us to evaluate PB changes such as duration, intensity, and timing of the different PAs performed over 24 hours, and relate them to metabolic health outcomes and their changes associated with the intervention in older adults. Figure 2 illustrates a bar plot with the average 24-hr predicted PA-distributions of all participants at the GOTO baseline accompanied by the ankle and wrist ENMO (black and white dash lines respectively).

**Figure 2:**
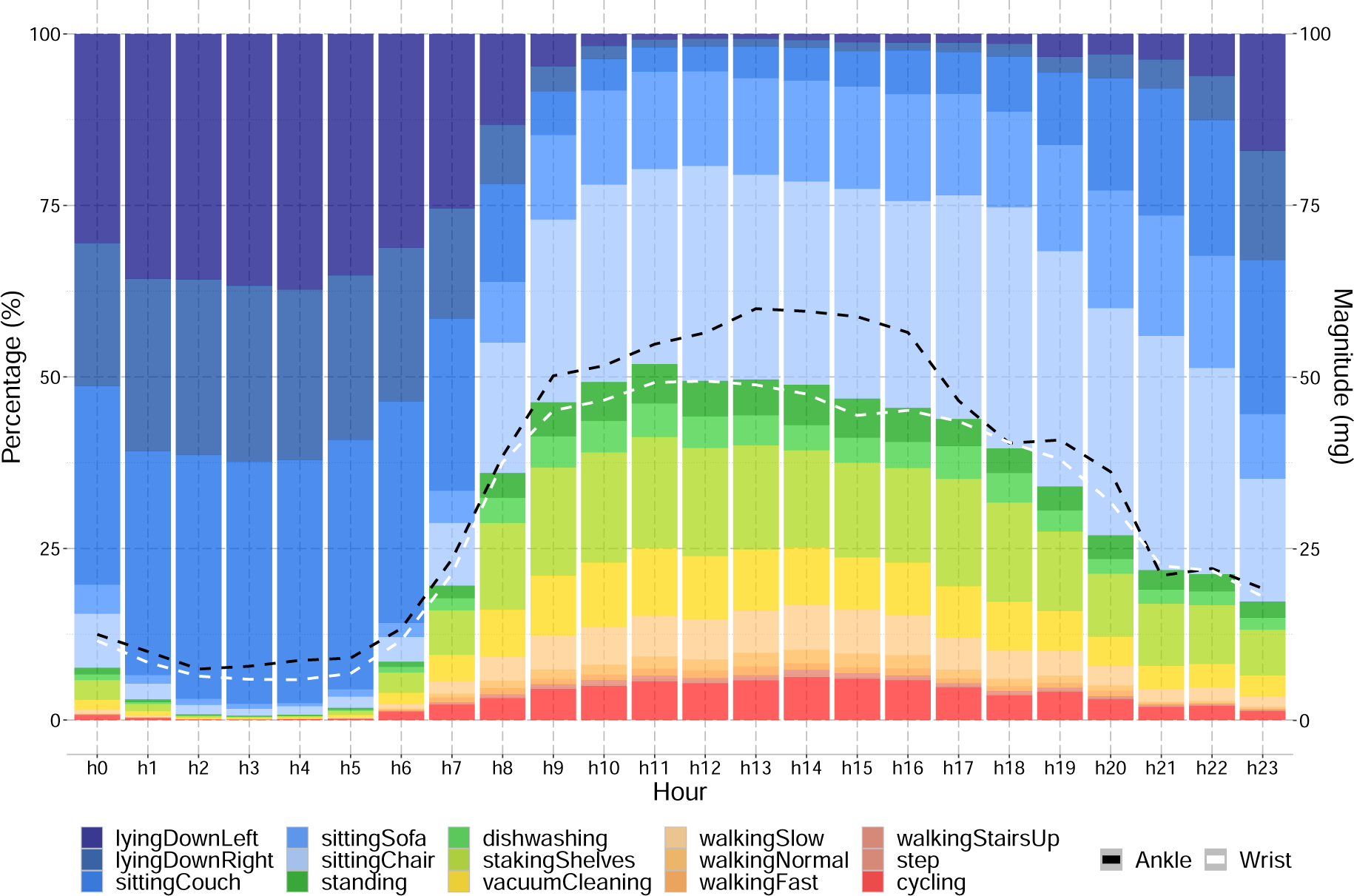
The average 24-hr PB distributions at baseline for all the participants in percentage per hour (left y-axis), along with the ankle and wrist ENMO (black and white dashed lines, respectively) in milligravity (right y-axis)

### 2.3 Data Inclusion

The analysis in this paper was carried out using a subset of 91 participants of the GOTO study for whom good quality data from both ankle and wrist, as well as all immune-metabolic biomarkers, were available, see Figure 3. The selection criteria in terms of accelerometer data included good quality data from both devices for at least one complete day (starting from 00:00 hours) both before and after the intervention. As a result, 51 participants were dropped based on the accelerometer criteria and another 22 based on the overlap with the selected immune-metabolic biomarkers.

**Figure 3:**
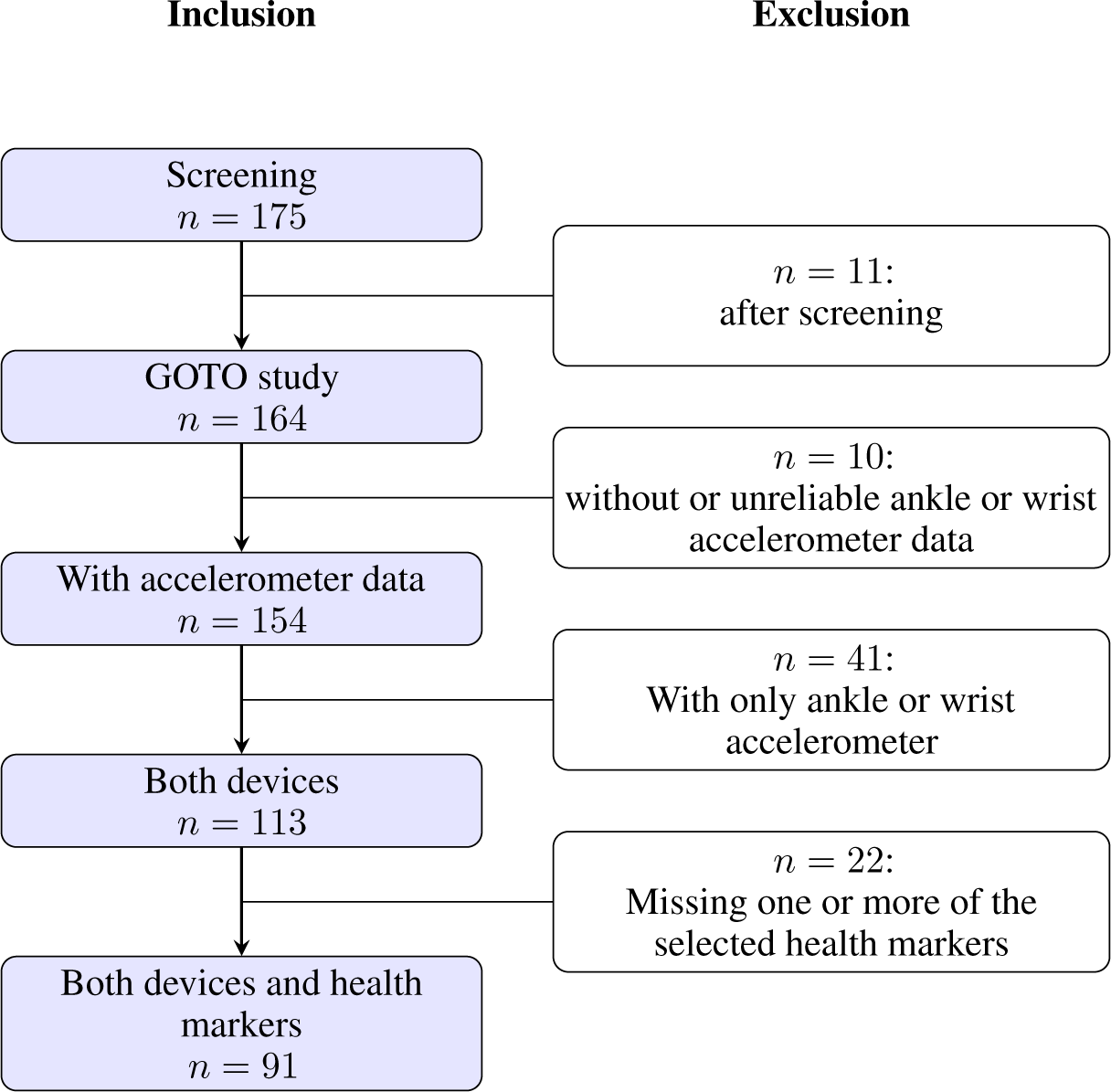
Data inclusion steps.

### 2.4 Study population

The current investigation aimed to explore the relationship between alterations in physical activity and various health parameters in a representative subset of the GOTO study (Table 1). The selected subgroup of the GOTO study has an average age of 62.6 years with 48.3% (N=44) of the participants being female. Table 2 demonstrates the levels of the various immune-metabolic parameters that were collected at baseline. There were no significant differences observed between our selected subset of the GOTO population and the overall GOTO sample, as reported in previous studies [29, 30]. Comparing male and female health parameters at baseline (see table 2), we can observe that most of the parameters are similar between men and women including BMI and inflammatory parameters like C-Reactive Protein (CRP). The main differences observed are linked to DEXA-estimated body/trunk fat percentages, with men having around 10 percent points lower fat percentages than women. Furthermore, male subjects had notably higher insulin levels than women, with a difference of about 2 mU/L.

**Table 1:** Description of data inclusion demographics compared to the total study population.

|  | All | Inclusion |
| --- | --- | --- |
| N | 164 | 91 |
| Sex (female %) | 49.4% | 48.3% |
| Age (years) | 63.0 ($\pm 5.7$) | 62.6 ($\pm 5.7$) |
| Weight (kg) | 79.2 ($\pm 9.8$) | 79.4 ($\pm 10.3$) |
| BMI (kg/m<sup>2</sup>) | 26.9 ($\pm 2.5$) | 26.8 ($\pm 2.5$) |

**Table 2:** Baseline levels of the selected health parameters (standard deviations)

| Baseline Mean (SD) | All (N=91) | Female (N=44) | Male (N=47) |
| --- | --- | --- | --- |
| Age in years | 62.6 ($\pm 5.7$) | 61.5 ($\pm 6.0$) | 63.7 ($\pm 5.4$) |
| Systolic Blood Pressure (mmHg) | 140.8 ($\pm 17.0$) | 139.4 ($\pm 18.0$) | 142.2 ($\pm 16.1$) |
| Diastolic Blood Pressure (mmHg) | 85.4 ($\pm 8.0$) | 85.0 ($\pm 9.1$) | 85.8 ($\pm 6.9$) |
| Weight (kg) | 79.4 ($\pm 10.3$) | 74.0 ($\pm 9.4$) | 84.4 ($\pm 8.5$) |
| Body Mass Index (kg/m<sup>2</sup>) | 26.8 ($\pm 2.5$) | 27.1 ($\pm 2.8$) | 26.6 ($\pm 2.2$) |
| Waist circumference (cm) | 96.1 ($\pm 8.3$) | 92.9 ($\pm 8.0$) | 99.1 ($\pm 7.6$) |
| Waist hip ratio (cm/cm) | 0.9 ($\pm 0.07$) | 0.9 ($\pm 0.05$) | 0.9 ($\pm 0.1$) |
| Whole body fat (%) | 32.0 ($\pm 7.4$) | 38.4 ($\pm 4.1$) | 26.1 ($\pm 4.2$) |
| Trunk fat (%) | 32.3 ($\pm 7.3$) | 37.2 ($\pm 5.7$) | 27.8 ($\pm 5.5$) |
| Glycerol (mmol/L) | 0.1 ($\pm 0.03$) | 0.1 ($\pm 0.03$) | 0.1 ($\pm 0.03$) |
| Glycoprotein acetyls (mmol/L) | 1.2 ($\pm 0.2$) | 1.2 ($\pm 0.1$) | 1.2 ($\pm 0.2$) |
| C-Reactive Protein (mg/L) | 1.8 ($\pm 2.7$) | 1.9 ($\pm 2.4$) | 1.7 ($\pm 2.9$) |
| Glucose (mmol/L) | 4.3 ($\pm 0.6$) | 4.2 ($\pm 0.5$) | 4.3 ($\pm 0.6$) |
| Insulin (mU/L) | 8.9 ($\pm 3.9$) | 7.9 ($\pm 3.8$) | 9.8 ($\pm 3.9$) |
| Framingham Risk Score | 13.7 ($\pm 2.2$) | 14.6 ($\pm 2.6$) | 13.0 ($\pm 1.6$) |
| Total Triglyceride (mmol/L) | 1.2 ($\pm 0.5$) | 1.0 ($\pm 0.4$) | 1.3 ($\pm 0.5$) |
| LDL-Cholesterol (mmol/L) | 1.9 ($\pm 0.5$) | 1.9 ($\pm 0.5$) | 1.9 ($\pm 0.5$) |
| LDL-Diameter (nm) | 23.6 ($\pm 0.1$) | 23.7 ($\pm 0.1$) | 23.6 ($\pm 0.1$) |
| HDL-Cholesterol (mmol/L) | 1.5 ($\pm 0.3$) | 1.7 ($\pm 0.3$) | 1.3 ($\pm 0.3$) |
| HDL-Diameter (nm) | 9.9 ($\pm 0.2$) | 10.0 ($\pm 0.2$) | 9.8 ($\pm 0.2$) |

### 2.5 Statistical analysis

We study the effect of physical behaviour in two ways: once looking at the cross-sectional effects at baseline (so ignoring the subsequent intervention), and once comparing the longitudinal changes in physical behaviour before and after to the changes in health before and after.

To study the cross-sectional effect of physical behaviour on the different health parameters at the *study baseline*, we performed sex-stratified multivariate linear regression, with the 19 different health parameters (see Table 2 for reference) as outcome and the 5 different PBs (two magnitude levels, and time spent on active, household, and sedentary PB) as the determinant, as presented in Equation 1. Adjustments were made for age at inclusion and inputs were standardised

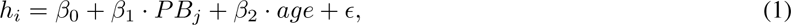

where *h_i_* represents one of the 19 different health parameters and *PB_j_* one of the different physical behaviours. As such, a total of 19 · 5 = 95 independent equations was produced (with *β*_0_*, β*_1_ and *β*_2_ not shared between equations).

The GOTO combined lifestyle intervention has effects on health parameters [29]. Here, to study the influence of *the longitudinal changes in physical behaviour resulting in changes in health parameters due to the intervention*, we performed sex-stratified multivariate linear mixed regression, with the selected 19 health parameters as an outcome and the different PBs (magnitude levels, and time spent on active, household, and sedentary PB) as the determinant, while adjusting for inclusion age (fixed effect) and taking the participants ID as a random effect:

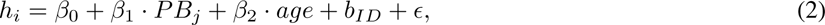

where *h_i_* again represents the 19 different health parameters as the outcome variable, *PB_j_* the 5 different physical behaviours as a predictor variable of interest, and *b_ID_* is the random-effects term associated with the participant’s ID. ^10^. For each pair (*h_i_, PB_j_*) of health parameter and PB, the following equation was fitted to the data To enhance our comprehension of the relationship between active and household physical behaviours and their impact on the health of individuals of both genders, we performed a deeper analysis. Specifically, we disassembled these behaviours into their underlying physical activity components and examined how *changes in longitudinal physical activities (PA) resulting from the intervention influenced alterations in health parameters*.

For active physical behaviour, the PA components were walking and cycling. Meanwhile, for household physical behaviour, the PA components included dishwashing, stacking shelves, and vacuum cleaning. This transformation is analogous to Equation 2, where the various physical behaviours (PBj) are substituted with different activity types (PAj), as illustrated in Equation 3:

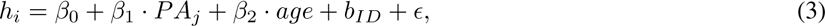

Here, *h_i_* represents the 19 different health parameters as the outcome variable, *PA_j_* denotes the various physical activities as predictor variables of interest, and *b_ID_* is the random-effects term associated with the participant’s ID. The model is stratified by sex and adjusted for age.

For our analyses, we used *R 4.1.2* (2021-11-01) and the R library *lme4* for Linear Mixed-Effects Models [32]. Furthermore, for plotting the different association heatmaps, we used *Python 3.10.2* and the packages *matplotlib*, and *seaborn* [33, 34].

## 3 Results

### 3.1 Physical behaviour at GOTO baseline resembles that of participants from the same age group in UKBiobank

To compare the estimated activity levels and physical behaviour in the older adults participating in the GOTO study with those in other population-based studies, we used data from the UKbiobank (UKB). This cohort includes 96.220 participants (more than 70.000 over 55 years old) participants who wore wrist accelerometers and reported both physical activity levels [35] and behaviour [13].

For a fair comparison and easier interpretation, we merged the 15 predicted activities to 5 as shown in Table 3. From this table, several general observations can be made regarding acceleration magnitude and the time spent on household activities, walking, and cycling. The accelerometer magnitudes reported by GOTO and UKB participants are similar^11^. Moreover, the GOTO participants (both male and female) spend on average approx. 75% of their time on sedentary activities like sitting or lying down (34.1% and 40.9% respectively in Table 3), 17% is spent on household activities and 8% on activities like walking or cycling (4.9% and 3.1% respectively). Compared to UKB [13], the GOTO participants spend a similar proportion of time sitting with 35.5% in GOTO and approx. 35% in UKB. However, GOTO participants spend more time lying down (40.9%) compared to UKB ones where lying down is reported as sleeping (approx. 37%). Similarly, the time spent on household activities (reported as mixed activity in UKB [13]) and walking are comparable. However, the GOTO participants report significantly more time spent cycling (3.1%) compared to those in UKB which report less than 1%, while they report an average of 5% of their day spent in a vehicle. This difference is expected given that the GOTO population is from the Netherlands, where cycling is more prevalent.

**Table 3:** Baseline physical activity percentages of the study population (standard deviation in brackets)

| Baseline Mean (SD) | All (N=91) | Female (N=44) | Male (N=47) |
| --- | --- | --- | --- |
| **Ankle magnitude in 24hr (mg)** | 35.4 ($\pm 12.4$) | 33.1 ($\pm 11.0$) | 35.6 ($\pm 12.8$) |
| **Wrist magnitude in 24hr (mg)** | 30.9 ($\pm 8.9$) | 29.6 ($\pm 8.2$) | 30.6 ($\pm 9.0$) |
| **Lying down (% in 24hr)** | 40.9 ($\pm 6.0$) | 41.9 ($\pm 5.8$) | 40.0 ($\pm 6.1$) |
| **Sitting/stand (% in 24hr)** | 34.1 ($\pm 6.6$) | 31.3 ($\pm 5.5$) | 36.8 ($\pm 6.5$) |
| **Household (% in 24hr)** | 16.9 ($\pm 4.4$) | 19.2 ($\pm 4.4$) | 14.8 ($\pm 3.3$) |
| **Walking (% in 24hr)** | 4.9 ($\pm 2.0$) | 4.7 ($\pm 1.6$) | 5.2 ($\pm 2.3$) |
| **Cycling (% in 24hr)** | 3.1 ($\pm 1.7$) | 2.9 ($\pm 1.4$) | 3.3 ($\pm 1.9$) |

Comparing the acceleration magnitudes for male and female participants, we observe in GOTO that on average, male participants show a higher mean magnitude for both ankle and wrist. This is something that was also observed in UKB [13] where only wrist magnitude is used. Women in GOTO spend a greater proportion of their time on household activities (19.2%) compared to men (14.8%). Additionally, men spend a greater proportion of their time sitting/standing (36.8%) compared to women (31.3%). For walking and cycling percentages we can observe that male participants spent slightly more time in these activities compared to the women. These findings are consistent with UKB [13], which also reported that women spend more time on mixed activities (reported as household activities in GOTO) compared to men, 18.9% and 14.5% respectively. Similarly, men in UKB [13] also spend more time sitting than women (34.6% and 37.3% respectively). However, in the GOTO study, women were found to spend less time sitting than those in UKB [13].

### 3.2 Men and women have physical behavioural differences throughout the day at baseline

To further investigate how physical behaviour varies across a 24-hour period, we explored three categories of behaviour: sedentary, household, and active. Sedentary behaviour includes the time spent sitting and lying down, active behaviour includes the time spent walking or cycling, and household behaviour includes the time spent on household activities. Table 4 presents the results of this comparison. Regardless of the time of day, participants spend the majority of their time in sedentary behaviour, with the highest percentage (97.6%) observed during the night and the lowest (52.6%) during the afternoon. In contrast, the afternoon is when participants are most active, with 15% of their time spent on active behaviour and 26.9% spent on household activities. When comparing the active behaviour of male and female participants, we observe that they both spent a similar amount of time on active activities during the afternoon, but the men spent slightly more time across the rest of the day, leading to a higher average percentage over the 24-hour period. On the other hand, there is a notable difference in household behaviour, with female participants spending an average of 5% more time on household activities compared to men. These findings are consistent with UKB [13].

**Table 4:** Baseline physical behaviour of the study population in percentages across the different times of the day and their standard deviation in brackets. The term active combines the time spent on walking and cycling, whereas sedentary combines lying down and sitting.

| Baseline Mean (SD) | All (N=91) | Female(N=44) | Male (N=47) |
| --- | --- | --- | --- |
| **Active (%)** |  |  |  |
| Night [00:00-05:59] | 0.5 ($\pm 0.5$) | 0.5 ($\pm 0.6$) | 0.5 ($\pm 0.5$) |
| Morning [06:00-11:59] | 9.5 ($\pm 4.7$) | 8.6 ($\pm 3.6$) | 10.2 ($\pm 5.5$) |
| Afternoon [12:00-17:59] | 15.0 ($\pm 5.7$) | 15.0 ($\pm 5.2$) | 15.0 ($\pm 6.1$) |
| Evening [18:00-23:59] | 7.2 ($\pm 3.9$) | 6.3 ($\pm 3.3$) | 7.9 ($\pm 4.2$) |
| 24 hours [00:00-23:59] | 8.0 ($\pm 2.9$) | 7.6 ($\pm 2.4$) | 8.4 ($\pm 3.3$) |
| **Household (%)** |  |  |  |
| Night [00:00-05:59] | 1.5 ($\pm 1.6$) | 1.6 ($\pm 1.6$) | 1.5 ($\pm 1.6$) |
| Morning [06:00-11:59] | 21.8 ($\pm 7.7$) | 25.1 ($\pm 8.5$) | 18.7 ($\pm 5.3$) |
| Afternoon [12:00-17:59] | 26.9 ($\pm 7.6$) | 30.1 ($\pm 7.4$) | 23.4 ($\pm 6.0$) |
| Evening [18:00-23:59] | 17.4 ($\pm 6.6$) | 19.4 ($\pm 7.3$) | 15.6 ($\pm 5.3$) |
| 24 hours [00:00-23:59] | 16.9 ($\pm 4.4$) | 19.2 ($\pm 4.4$) | 14.8 ($\pm 3.3$) |
| **Sedentary (%)** |  |  |  |
| Night [00:00-05:59] | 97.6 ($\pm 2.2$) | 97.7 ($\pm 2.1$) | 97.6 ($\pm 2.2$) |
| Morning [06:00-11:59] | 64.9 ($\pm 10.5$) | 63.1 ($\pm 10.7$) | 66.6 ($\pm 10.1$) |
| Afternoon [12:00-17:59] | 52.6 ($\pm 10.6$) | 49.8 ($\pm 11.1$) | 55.2 ($\pm 9.5$) |
| Evening [18:00-23:59] | 72.2 ($\pm 9.1$) | 71.8 ($\pm 9.9$) | 72.6 ($\pm 8.3$) |
| 24 hours [00:00-23:59] | 71.8 ($\pm 5.8$) | 70.6 ($\pm 6.0$) | 73.0 ($\pm 5.4$) |

To examine a potential difference in PB between men and women, we conducted Kolmogorov-Smirnov (K-S) tests on the 24-hour averages as shown in Table 3, considering the non-normal distributions of our data. Notably, we observed that men spent less time than women lying down (*p* = 0.066) and performing household activities (*p* = 1.0 · 10*^−^*^5^). We furthermore observed that men spent more time sitting/standing (*p* = 1.6 · 10*^−^*^3^) than women. However, no significant differences were found in other activities, including ankle and wrist 24-hour averages (refer to Table 6 in the Appendix).

Furthermore, Figure 4 provides insights into the activity patterns of male and female participants. This figure illustrates the average percentage point differences in activity duration per hour (from h0 to h23) for each activity type (represented by the stacked colors) between women and men. For example, between 08:00 and 08:59 (h8), women predominantly engaged in household tasks, notably stacking shelves (shown in light green), whereas men were more inclined to sit, particularly on chairs. At the same time, we don’t observe any big differences in the higher-intensity activities e.g. walking and cycling (depicted in orange and red color).

**Figure 4:**
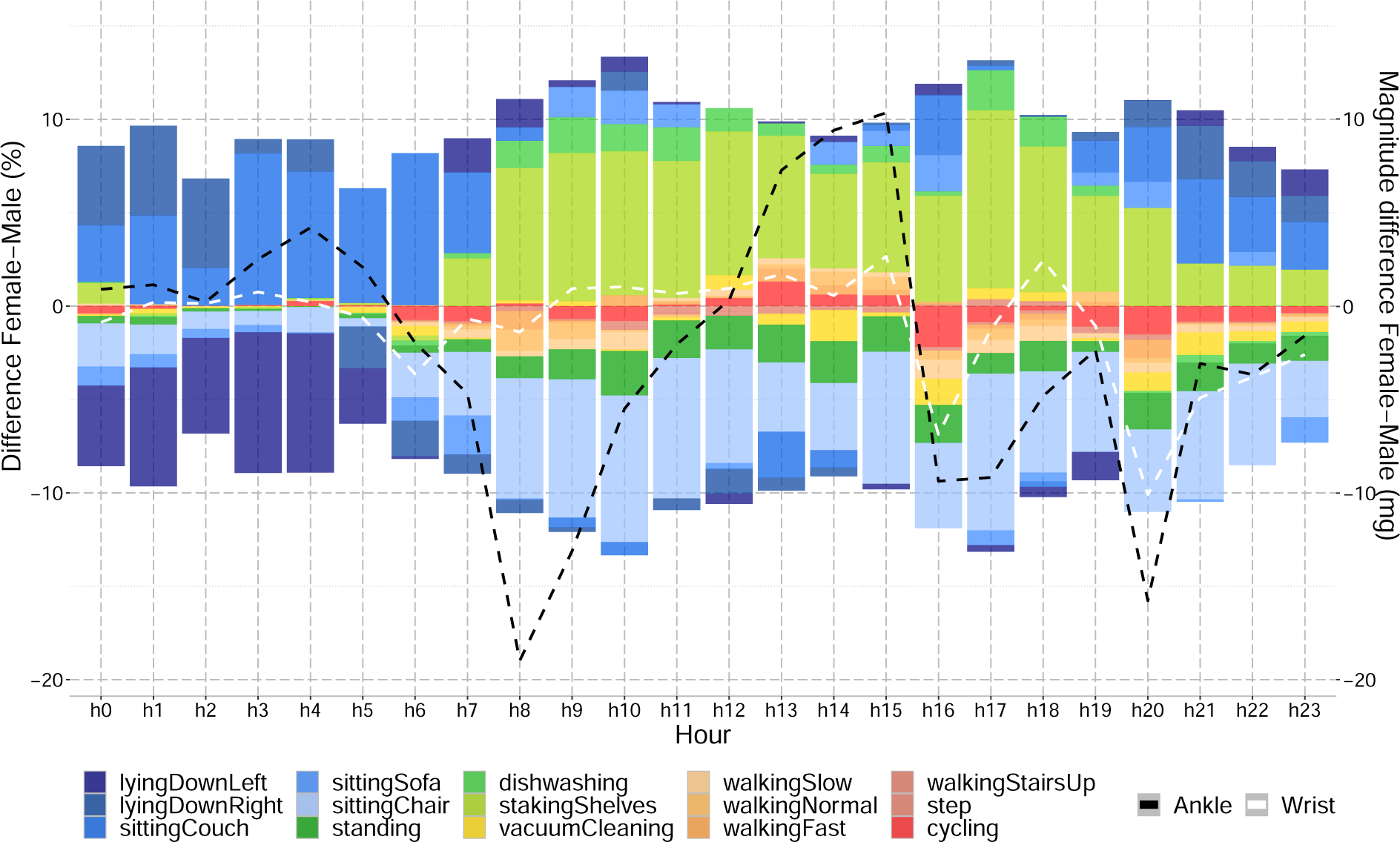
Differences of activities between female and male participants at baseline over 24hrs. Positive values indicate higher percentages or magnitudes (mg) for women and negative indicate higher for men. The left y-axis presents the percentage points difference and the right y-axis the activity magnitude scale.

Overall, on average, male participants dedicate more time to sedentary activities (indicated by blue colours) throughout the day, while female participants tend to spend more time engaging in household activities (depicted by light greens). However, a notable contrast is observed in the distribution of active behaviours (represented by red colours) and standing (depicted by darker green colour). Male participants allocate a greater proportion of their day to active behaviours and standing compared to women.

Moreover, an analysis of ankle and wrist magnitudes indicates that male participants are more involved in higher ankle magnitude activities (represented by the black dashed line in Figure 4). This trend is prominent during various periods throughout the day. For wrist activities, the higher magnitude trend is predominantly observed during the evening hours (as evident from the white dashed line).

We proceeded to investigate the significance of differences in the daily patterns between males and females, as presented in Table 4 and Figure 4, using ANOVA testing with Sphericity Corrections. The ANOVA results (see ANOVA equation 4 in the Appendix) confirmed that household activity exhibits significant differences between men and women across various parts of the day (night, morning, afternoon, and evening). Additionally, when examining the data at an hourly level, we found that the activity patterns of men and women significantly differ for each hour, indicating a notable impact of time of day on physical behaviour and ankle/wrist magnitudes. The Sphericity Corrections further validate the ANOVA findings. Specifically, the ANOVA results indicate that the changes in patterns per hour, and per gender, are significant for all variables except wrist magnitude.

Also, the Sphericity Corrections reinforce the significance of the interaction between gender and time, as evidenced by low p-values for the Greenhouse-Geisser and Huynh-Feldt corrections. After the correction, the significance of the active physical behaviour (PB) percentage was attenuated (*p* = 0.081, refer to Tables 9 and 10 in the Appendix). Overall, the results underscore the influence of gender and time on activity patterns and ankle/wrist magnitudes, shedding light on important behavioural differences between men and women throughout the day.

### 3.3 Higher levels of PB at baseline correlate with a healthier immune-metabolic profile

The cross-sectional relationships between activity magnitudes (either ankle or wrist ENMO) and various health markers, as well as their associations with physical activity behaviours at the GOTO baseline, are illustrated in a heatmap in Figure 5. These associations are modeled using linear regression, stratified by sex, and adjusted for age, as shown in Equation 1. For interpretation, lower levels of all health markers indicate better health, with the exception of HDL-cholesterol and cholesterol diameter parameters, for which the opposite is true (refer to signs next to parameters name in Figure 5).

**Figure 5:**
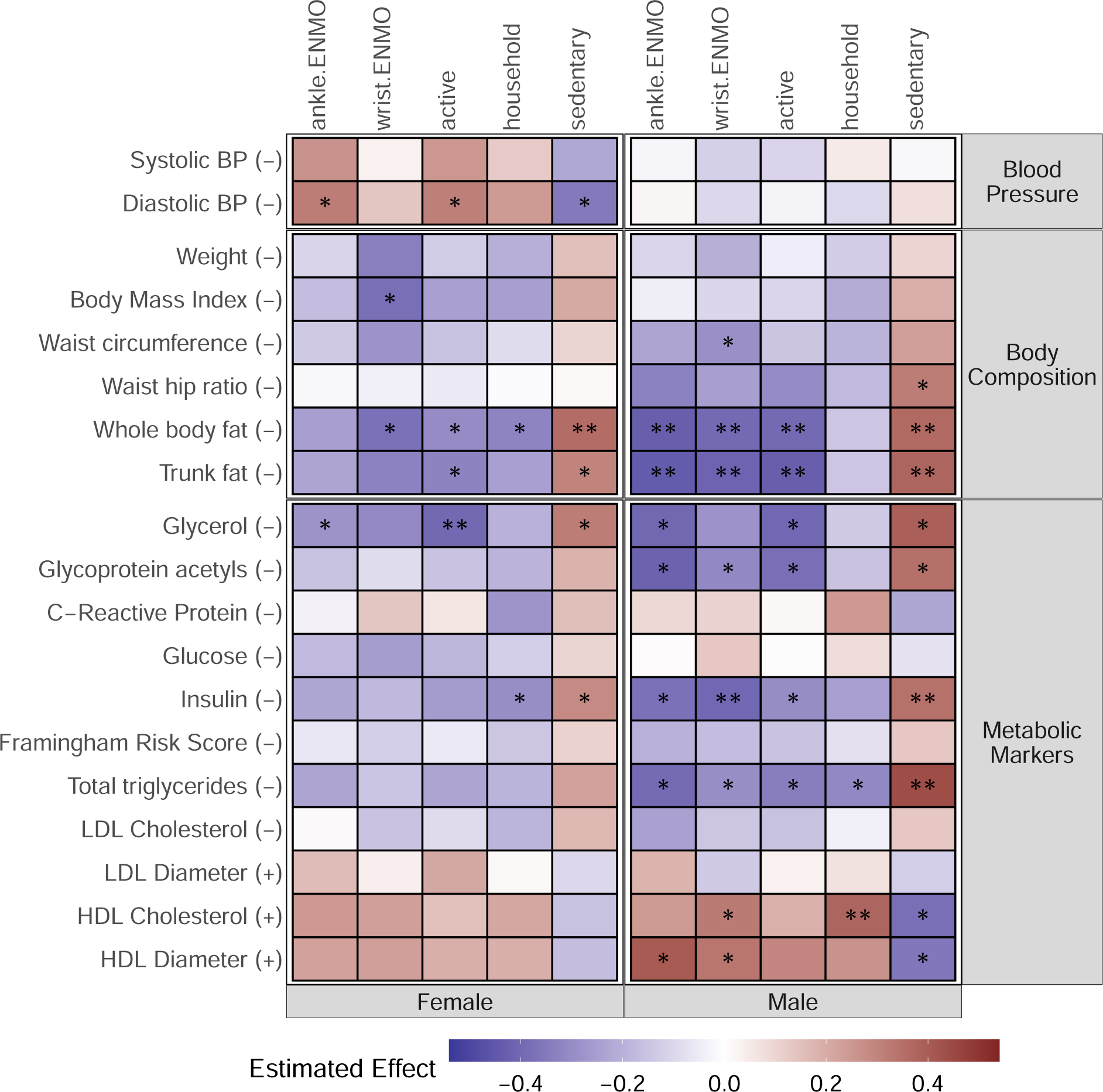
Associations of physical behaviour with health markers at baseline for female (left) and male (right) participants. Darker blue indicates lower levels of health markers, while darker red has higher levels. Stars indicate significant associations with one asterisk *p <* 0.05, two asterisks *p <* 0.01, and three asterisks *p <* 0.001. Low levels of markers are all an indication of better health as indicated by (*−*) except for the LDL, HDL diameter parameters, and HDL cholesterol for which the reverse is true as indicated by (+) next to them.

The results demonstrate the extent to which health variance at baseline is connected to pre-existing differences in physical behaviour (PB). Specifically, parameters of overall body composition, such as BMI and waist circumference, as well as more refined DEXA-fat parameters and glycerol, are associated with PB in both genders and contrasting effects of active and sedentary PB are observed. There are some striking sex differences to be observed from Figure 5. The association of all PB behavioural parameters with triglycerides and HDL-cholesterol features are much stronger in men than in women whereas the serum levels are the same in both sexes (see Table 2). Serum insulin is higher in men and associates with PB parameters whereas in women only with household activities. Interestingly, the classical clinical marker indicating inflammation (circulating C-Reactive Protein levels) is not associated with PB while a second relevant inflammatory marker, circulating levels of glycoprotein acetyls, is associated with activity in men and not in women. In women, blood pressure parameters exhibit a correlation with PB, although this association is somewhat unexpected and can be attributed to the relationship between blood pressure parameters and BMI (refer to Figure **??** in the supplementary material for further details). Furthermore, in women, household activities do not correlate with many health parameters except for whole body fat and insulin as observed in women.

We can interpret the standardized results shown in Figure 5 by converting the effect estimates back to the original scale of the variables. The unstandardized LM-estimates can be found in the Appendix (refer to Table 11 for male and Table 12 for women estimates). This allows us to examine the relationship between the magnitude or time spent on activity levels and health benefits. For instance, we find that an additional minute per day spent on active activities was strongly associated with 0.15% lower whole-body fat for male participants and 0.11% for women. Conversely, an extra minute in sedentary activities was strongly associated with 0.26% more whole body fat for women and 0.27% for men. Furthermore, we observe that, on average, an additional 1 mg of ankle magnitude per day^12^ in men was strongly associated among others with a 0.68% lower whole body fat, 0.69% lower trunk fat, 1.03 less mmol/L of insulin, and 20.05 nm bigger HDL-Diameter. Similarly, an extra mg of wrist magnitude was strongly associated with a 0.47% lower whole body fat, 0.49% lower trunk fat, 0.84 less mU/L of insulin, and 12.7 nm bigger HDL-Diameter. For women, an extra 1 mg of wrist magnitude per day^13^ was strongly associated with a 0.45% less whole body fat and 1.33 lower body mass index, while an extra 1 mg of the ankle was strongly associated with 113 less mmol/L of Glycerol.

### 3.4 Increase in activity level and physical behaviour due to lifestyle intervention

We consecutively explore whether the lifestyle intervention protocol had an effect on physical behaviour (Table 5). On average, the active time of all participants increased by 4.3% overall. However, this increase is mostly explained by an increase amongst the female participants, with an 8.2% increase, compared to a modest 0.6% increase among male participants, although with a high standard deviation of 22.8% (suggesting that some participants didn’t change their behaviour whereas others did substantially). Women seemed to mainly increase the activity of the upper limbs and men of lower limbs, The household time remained mostly unchanged with a mean difference of 0.7%. In contrast, although the average sedentary time appeared to remain unchanged, there was a small increase of 1.2% in male participants.

**Table 5:**
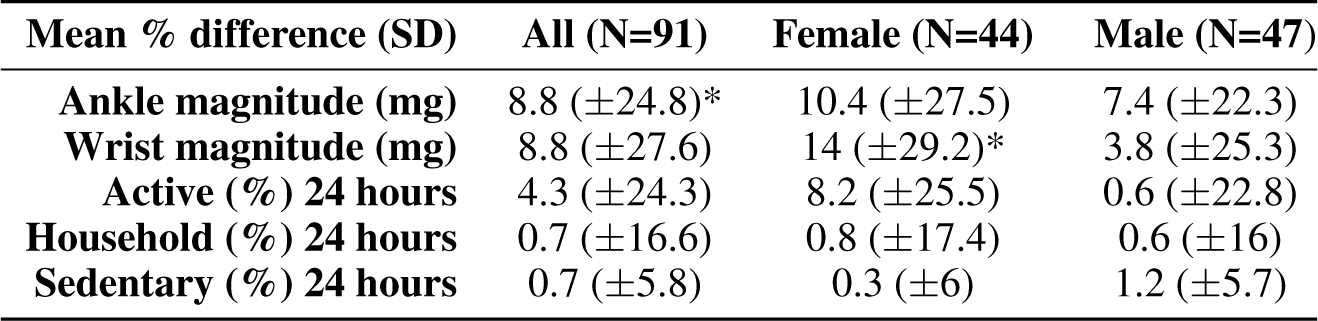
Percentage changes of physical behaviour between before and after the intervention where star indicates a significant difference.

### 3.5 Increasing the time of being active is beneficial for the physical health of older people

The relationships between longitudinal changes in average magnitudes or time spent on specific physical behaviours and the changes in health markers due to the intervention are displayed in a heatmap in Figure 6. This heatmap was generated by modeling the relationships using a linear mixed model (as described in Equation 2), stratified by sex and adjusted for age.

**Figure 6:**
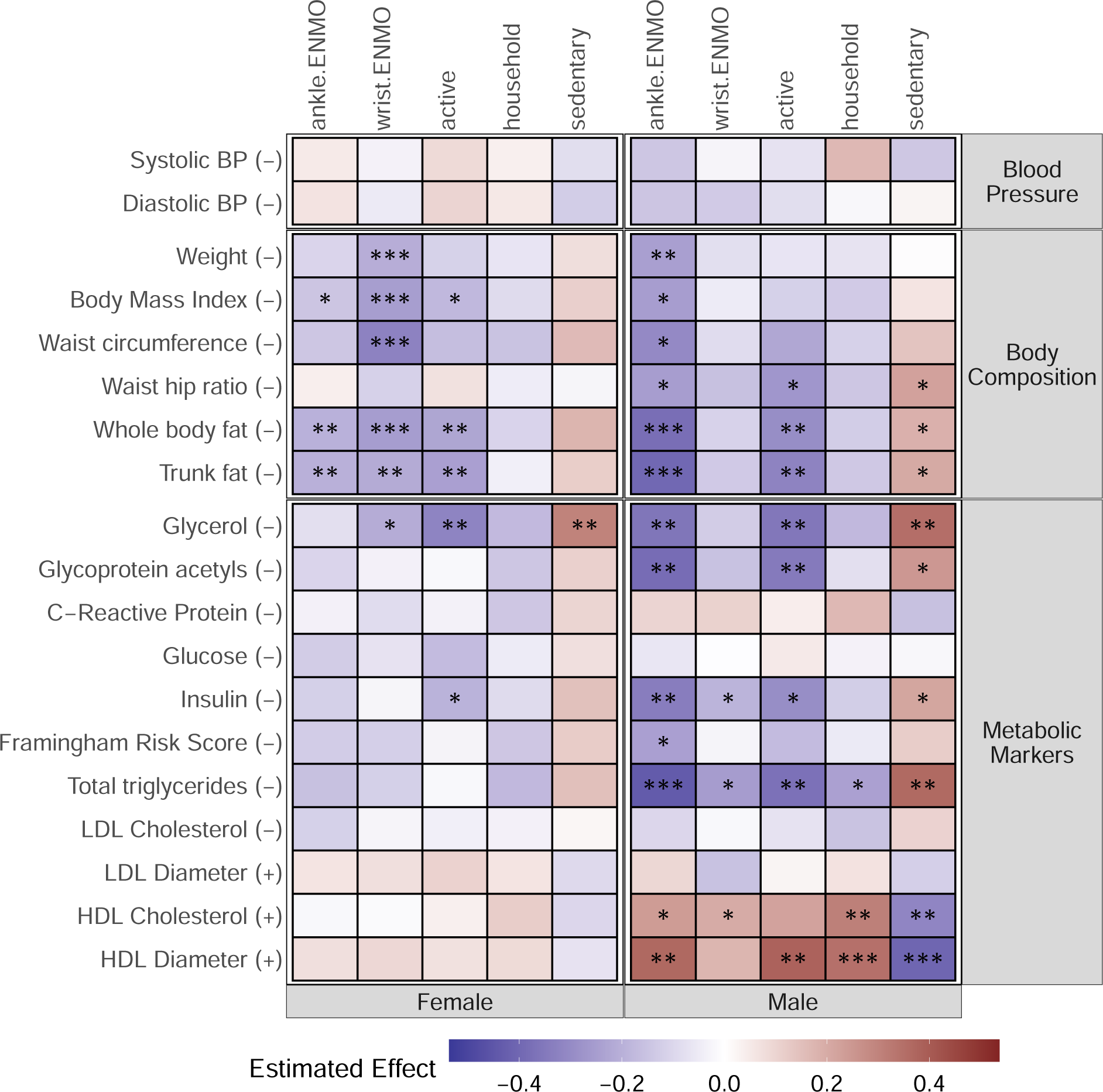
Associations of physical behaviour change with the change of health markers levels after the intervention for female (top) and male (bottom) participants. Darker blue indicates lower levels of health markers, while darker red has higher levels. Stars indicate significant associations with one star *p <* 0.05, two stars *p <* 0.01, and three stars *p <* 0.001. Low levels of markers are all an indication of beneficial health change as indicated by (*−*) except for the LDL, HDL diameter parameters, and HDL cholesterol for which the reverse is true as indicated by (+) next to them.

For both sexes, an increase in PB is associated with beneficial changes in body composition, refined DEXA-based fat parameters, glycerol, and insulin, while an increase in sedentary behaviour exhibits contrasting effects. In detail, these benefits were more pronounced in men and especially driven by lower limb activity (see ankle ENMO magnitude). On the other hand, it was striking that changes in PB for women provided much fewer health benefits as represented by the health parameters. For men, active PB changes were related to beneficial changes in total triglycerides, HDL-cholesterol features, and the inflammatory marker glycoprotein acetyls levels (but not C-Reactive Protein levels). Interestingly, a slight increase in household activity for men was gained by the intervention beneficially affecting total triglyceride levels and HDL parameters. Collectively, these results point to a positive association between changes in PB during the intervention and health improvements illustrated by the same set of parameters and their direction as observed for baseline relations, including the more prominent beneficial effects in men.

We can interpret the standardized results presented in Figure 6 by converting the effect estimates of the LMM model back to the original scale of the variables. The unstandardized LMM-estimates can be found in the Appendix (refer to Table 13 for male and Table 14 for women estimates). This allows us to examine the associations between the magnitude or time spent on activity levels and health benefits after the intervention. For example, we found that, on average, 1 mg increase in activity levels per day resulted in a decrease of 1.16 kg/m^2^ in BMI for men and 0.67 kg/m^2^ for women after the intervention. Similarly, a 1 minute increase in time spent on active activities per day was strongly associated with a 0.11% decrease in whole body fat for men and a 0.08% decrease for women after the intervention. In addition, among men, we observed that a 1 minute increase in active activities per day after the intervention was strongly associated with beneficial increases in HDL-Diameter by 4.46 nm and a decrease in insulin by 0.4 mU/L. On the other hand, a 1 minute increase in sedentary time after the intervention was strongly associated with a decrease in HDL-Diameter by 9.38 nm and an increase in insulin by 0.28 mU/L which both are non-beneficial.

Furthermore, we observe that, on average, increasing 1 mg of **ankle** magnitude per day in men was strongly associated, among others, with 0.28 kg less body weight, 0.6% less whole body and trunk fat, 146.5 less mmol/L of Glycerol, 27.0 less mmol/L of Glycoprotein acetyls, 0.93 less mU/L of insulin, 10.4 less mmol/L of total triglyceride, and 18.1 nm bigger HDL-Diameter. In contrast, an increase of 1 mg of **wrist** magnitude after the intervention in men is only associated with 0.4 less mU/L of insulin, 4.6 less mmol/L of total triglyceride, and 5.8 less mmol/L HDL-Cholesterol. For women, on the other hand, most associations are observed from wrist magnitude, with an increase of 1 mg being strongly linked to 0.18 less kg of body weight, 0.88 lower BMI, and 0.3% less body and trunk fat.

### 3.6 Increasing time in more intense activity types is beneficial for the physical health of older people

To gain a deeper understanding of the relation between physical behaviours and their effects on health for both sexes, we decomposed the two more intense physical behaviours (active and household) into their physical activity (PA) components. For active physical behaviour, the PA components include walking and cycling, while for household physical behaviour, the PA components consist of dishwashing, stacking shelves, and vacuum cleaning. Figure 7 presents the associations’ heatmap for the different PAs within these two behaviours. The heatmap was generated by modeling the relationships using Equation 3.

**Figure 7:**
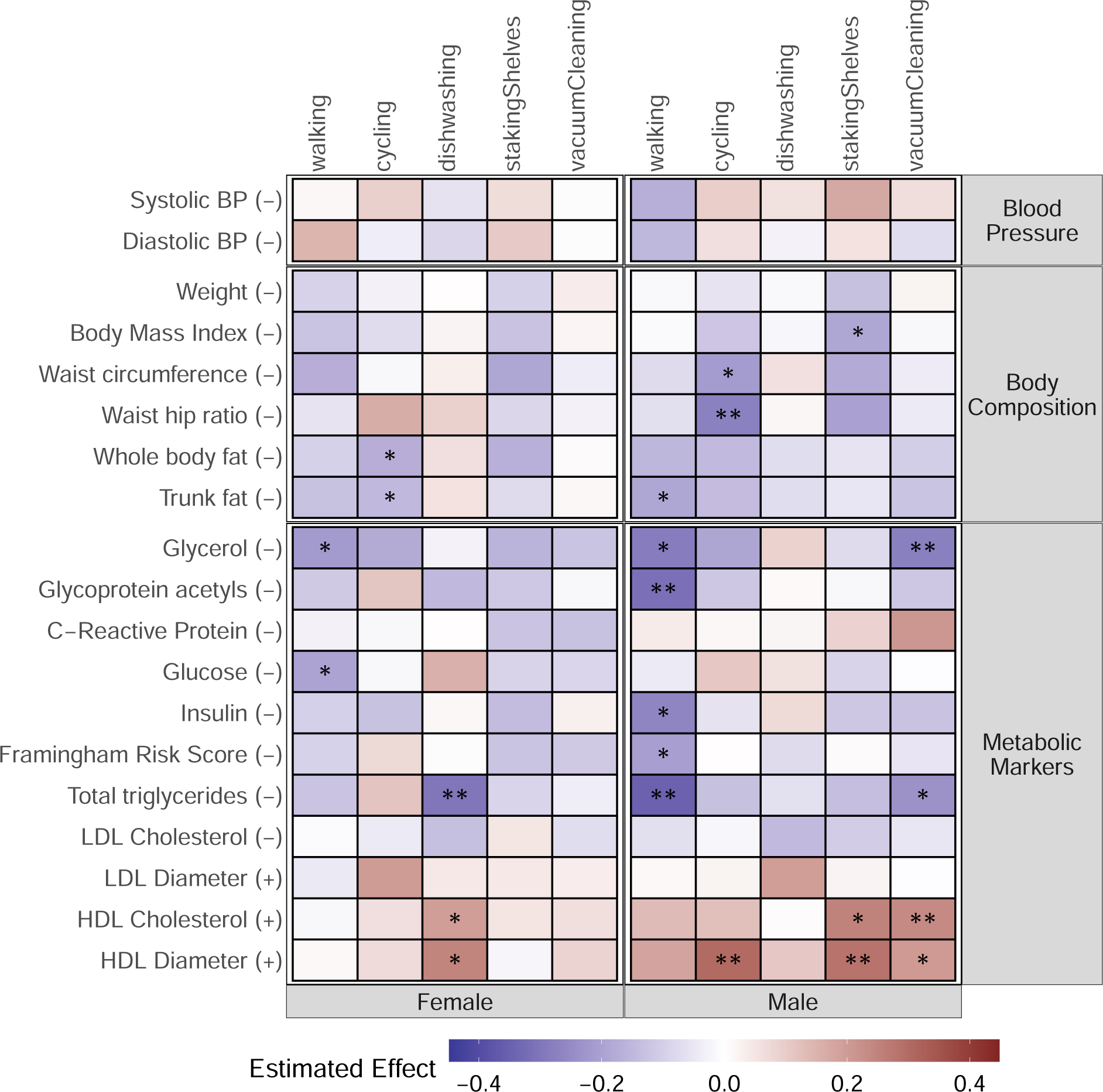
Associations of active and household physical activities change with the change of health markers levels after the intervention for female (top) and male (bottom) participants. Darker blue indicates lower levels of health markers, while darker red has higher levels. Stars indicate significant associations with one star *p <* 0.05, two stars *p <* 0.01, and three stars *p <* 0.001. Low levels of markers are all an indication of beneficial health change as indicated by (*−*) except for the LDL, HDL diameter parameters, and HDL cholesterol for which the reverse is true as indicated by (+) next to them.

The results suggest that increasing time spent on PA types driven by lower limbs, such as walking and cycling, is associated with beneficial changes in body composition, refined DEXA-based fat parameters, glycerol, and insulin for both sexes. When inspecting sexes separately we observed overall, stronger effects for men. Specifically, for men, increased time spent on vacuum cleaning and stacking shelves PAs, which both involve upper and lower limb activity, was strongly associated with beneficial changes in HDL-cholesterol, glycerol, and triglycerides. The unstandardized LMM-estimates for activities can be found in the Appendix (refer to Table 15 for male and Table 16 for women estimates). In detail, a 1 minute increase of vacuum cleaning on average per day is strongly associated with a decrease of 0.74 mmol/L in triglycerides levels and 14.77 mmol/L of glycerol and at the same time with an increase of 1.18 mmol/L and 1.32 nm of HDL-Cholesterol and HDL-Diameter respectively. Conversely, for women, a strong beneficial effect was observed with increased time spent on dishwashing, with an increase of a 1 minute spent being associated with a decrease of 0.77 mmol/L in triglycerides levels and at the same time an increase of 0.8 mmol/L and 1.31 nm of HDL-Cholesterol and HDL-Diameter respectively. These findings indicate a positive association between health improvement and increased time in higher-intensity PA types.

## 4 Discussion

We studied accelerometer-based physical activity (PA) and behaviour (PB) in a 13-weeks combined lifestyle intervention study in older adults, the GOTO study. We observed that men and women have different physical behaviour patterns throughout the day: as compared to women, men spend more time sedentary while having more episodes of intense activities. In cross-sectional analyses of the baseline data, high levels of activity and more time spent while walking and cycling are associated with beneficial levels of immune-metabolic markers. These effects are, as compared to women, much more pronounced in men. Moreover, an increase in PB due to the GOTO lifestyle intervention, is associated with beneficial changes in body composition in both men and women. In addition, an increase in PB is associated with beneficial levels of immune-metabolic markers in men, and not in women. Finally, our study showcases the effectiveness of using machine learning models like LARA to observe the effects of different physical behaviours in intervention studies. In conclusion, our findings strongly support the positive impact of increased physical behaviour on immune-metabolic health in older adults. These results underscore the importance of encouraging and promoting physical activity among older individuals to enhance their overall immune-metabolic health. Our study highlights the need to further investigate the specific PB changes required for women to achieve health benefits comparable to men.

We demonstrate that even a mild, short combined lifestyle change in ages above 60 contributes to health improvement exemplified by body composition and a profile of serum parameters associated with increased vitality. By leveraging activity recognition models like LARA and focusing even more closely on sex-specific behaviours and benefits, we take a step closer to providing straightforward lifestyle guidelines, as they allow for a comprehensive and nuanced understanding of the effectiveness of individual physical behaviours at older ages.

The sex differences we observed correspond at baseline and in response to the intervention. Despite women exhibiting more overall movement throughout the day, men derive greater benefits exemplified in serum profiles (decrease in glycoprotein acetyls, and triglycerides; increase in HDL-related parameters). This seems most likely due to the intensity of their activity but it may be also due to differences in diet, muscle composition, hormonal status, or metabolic flexibility. Strikingly, men showed a more significant health benefit associated with activities like vacuuming and walking compared to women, even though they spent more time being sedentary, as depicted in Figure 7. These sex differences in the effect size of PB on health parameters necessitate a more in-depth investigation in subsequent studies, addressing the question: "Which PB is most beneficial for whom?" especially when evaluating health improvements via serum indicators.

The GOTO study has several limitations. First, it examines the effects of a combined lifestyle intervention, encompassing at the same time increased PB and decreased caloric intake. In the current study design, it is hard to disentangle the separate effects. It may for example be that the people that increased their PB the most also decreased their caloric intake the most. Unfortunately, the reduction of caloric intake has not been registered quantitatively. Second, the GOTO study has a limited sample size, which even further decreases when we feel the need for stratified analyses. Third, by using activity recognition models we rely on behaviours labeled by LARA which may hide detailed differences in behaviour. However, we feel that the use of LARA is also a strength of the current study because it gives good insight into the physical behaviour of older adults. The GOTO study provides firm evidence for a widespread health improvement to be expected from longer-term lifestyle changes, i.e. longer than 13 weeks as in the GOTO protocol. In addition, it indicates through the sex-specific effects that studying personal effects more intensely will deliver evidence-based recommendations for effective changes in physical behaviour in older adults. We expect these findings to support designing community-based programs that promote physical activity and improve metabolic health outcomes in older adults.

## 5 Conclusion

Concluding, the results of the GOTO study provide significant insights into the impact of mild interventions in older adults and emphasize the importance of physical behaviour for boosting and maintaining healthy body composition and serum health profiles of parameters involved in glucose and lipid metabolism, and inflammation. These findings highlight the benefits of improving physical behaviour and underscore the need to combat sedentary lifestyles and associated metabolic diseases in older adults. The study particularly contributes to our understanding of sex differences in the choice, frequency, and intensity of physical behaviours and emphasizes the large variation in effect on health improvement when recorded in depth by DEXA scans and serum assays. The study warrants more research to explore by which physical behaviour women gain health benefits comparable to men and how this can be maximized for older adults of both sexes.

## Data Availability

All data produced in the present study are available upon reasonable request to the authors

## Acknowledgements

The authors express their gratitude to all participants of the GOTO study who did their very best to adhere to the intervention guidelines and underwent all measurements. FUNDING INFORMATION: This work was funded by the Netherlands Consortium for Healthy Ageing (NWO grant 050-060-810), the framework of the BBMRI Metabolomics Consortium funded by BBMRI-NL (NWO 184.021.007 and 184.033.111) and the VOILA Consortium (ZonMw 457001001). The funding agencies had no role in the design and conduct of the study; collection, management, analysis, and interpretation of the data; and preparation, review, or approval of the manuscript.

## APPENDIX

**Table 6:** Baseline Activity - Significance Test Results comparing men and women ANOVA equation:

| Baseline-Activity | p-value | Significance |
| --- | --- | --- |
| Ankle magnitude in 24hr (mg) | 0.7087 |  |
| Wrist magnitude in 24hr (mg) | 0.3781 |  |
| Lying down (% in 24hr) | 0.0657 |  |
| Sitting/stand (% in 24hr) | 0.0016 | * |
| Household (% in 24hr) | 0.00001 | * |
| Walking (% in 24hr) | 0.6417 |  |
| Cycling (% in 24hr) | 0.7099 |  |

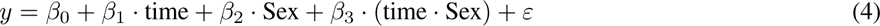

**Table 7:** ANOVA results comparing men and women per part of the day (night, morning, afternoon, evening) at baseline.

| Activity | Effect | F | p | p < .05 | ges |
| --- | --- | --- | --- | --- | --- |
| Active (%) | Sex | 1.718 | 0.193 |  | 0.009 |
| Active (%) | time | 276.661 | 1.35e-81 | * | 0.614 |
| Active (%) | Sex:time | 1.621 | 0.185 |  | 0.009 |
| Household (%) | Sex | 29.380 | 5.04e-07 | * | 0.125 |
| Household (%) | time | 426.681 | 1.65e-101 | * | 0.731 |
| Household (%) | Sex:time | 9.261 | 7.57e-06 | * | 0.056 |
| Sedentary (%) | Sex | 3.986 | 0.049 | * | 0.019 |
| Sedentary (%) | time | 579.567 | 1.49e-116 | * | 0.787 |
| Sedentary (%) | Sex:time | 2.569 | 0.055 |  | 0.016 |
| Ankle magnitude (mg) | Sex | 0.923 | 0.339 |  | 0.005 |
| Ankle magnitude (mg) | time | 192.865 | 1.58e-66 | * | 0.529 |
| Ankle magnitude (mg) | Sex:time | 2.551 | 0.056 |  | 0.015 |
| Wrist magnitude (mg) | Sex | 0.346 | 0.558 |  | 0.002 |
| Wrist magnitude (mg) | time | 276.617 | 1.37e-81 | * | 0.597 |
| Wrist magnitude (mg) | Sex:time | 0.589 | 0.623 |  | 0.003 |

**Table 8:** Sphericity Corrections comparing men and women per part of the day (night, morning, afternoon, evening) at baseline.

| Activity | Effect | GGe | p[GG] | p[GG] < .05 | HFe | p[HF] | p[HF] < .05 |
| --- | --- | --- | --- | --- | --- | --- | --- |
| Active (%) | time | 0.869 | 2.900e-71 | * | 0.897 | 1.640e-73 | * |
| Active (%) | Sex:time | 0.869 | 1.910e-01 |  | 0.897 | 1.900e-01 |  |
| Household (%) | time | 0.877 | 2.420e-89 | * | 0.906 | 3.250e-92 | * |
| Household (%) | Sex:time | 0.877 | 2.270e-05 | * | 0.906 | 1.750e-05 | * |
| Sedentary (%) | time | 0.912 | 1.480e-106 | * | 0.944 | 3.640e-110 | * |
| Sedentary (%) | Sex:time | 0.912 | 6.000e-02 |  | 0.944 | 5.800e-02 |  |
| Ankle magnitude (mg) | time | 0.933 | 2.850e-62 | * | 0.967 | 2.120e-64 | * |
| Ankle magnitude (mg) | Sex:time | 0.933 | 6.000e-02 |  | 0.967 | 5.800e-02 |  |
| Wrist magnitude (mg) | time | 0.969 | 3.990e-79 | * | 1.005 | 1.370e-81 | * |
| Wrist magnitude (mg) | Sex:time | 0.969 | 6.170e-01 |  | 1.005 | 6.230e-01 |  |

**Table 9:** ANOVA results comparing men and women per hour of the day at baseline.

| score | Effect | DFn | DFd | F | p | p< .05 | ges |
| --- | --- | --- | --- | --- | --- | --- | --- |
| Active (%) | Sex | 1 | 89 | 1.718 | 0.193 |  | 0.004 |
| Active (%) | time | 23 | 2047 | 103.52 | 0.00 | * | 0.48 |
| Active (%) | Sex:time | 23 | 2047 | 1.657 | 0.026 | * | 0.014 |
| Household (%) | Sex | 1 | 89 | 29.381 | 0.00 | * | 0.068 |
| Household (%) | time | 23 | 2047 | 216.51 | 0.00 | * | 0.65 |
| Household (%) | Sex:time | 23 | 2047 | 7.390 | 0.00 | * | 0.061 |
| Sedentary (%) | Sex | 1 | 89 | 3.986 | 0.049 | * | 0.01 |
| Sedentary (%) | time | 23 | 2047 | 282.59 | 0.00 | * | 0.71 |
| Sedentary (%) | Sex:time | 23 | 2047 | 2.622 | 0.000 | * | 0.022 |
| Ankle magnitude (mg) | Sex | 1 | 89 | 0.923 | 0.339 |  | 0.002 |
| Ankle magnitude (mg) | time | 23 | 2047 | 66.37 | 0.000 | * | 0.37 |
| Ankle magnitude (mg) | Sex:time | 23 | 2047 | 1.973 | 0.004 | * | 0.017 |
| Wrist magnitude (mg) | Sex | 1 | 89 | 0.346 | 0.558 |  | 0.001 |
| Wrist magnitude (mg) | time | 23 | 2047 | 128.36 | 0.00 | * | 0.51 |
| Wrist magnitude (mg) | Sex:time | 23 | 2047 | 1.003 | 0.457 |  | 0.008 |

**Table 10:** Sphericity Corrections comparing men and women per hour of the day at baseline.

| score | Effect | GGe | p[GG] | p[GG]< .05 | HFe | p[HF] | p[HF]< .05 |
| --- | --- | --- | --- | --- | --- | --- | --- |
| Active (%) | time | 0.411 | 0.000 | * | 0.464 | 0.000 | * |
| Active (%) | Sex:time | 0.411 | 0.091 |  | 0.464 | 0.081 |  |
| Household (%) | time | 0.370 | 0.000 | * | 0.413 | 0.000 | * |
| Household (%) | Sex:time | 0.370 | 0.000 | * | 0.413 | 0.000 | * |
| Sedentary (%) | time | 0.380 | 0.000 | * | 0.425 | 0.000 | * |
| Sedentary (%) | Sex:time | 0.380 | 0.006 | * | 0.425 | 0.004 | * |
| Ankle magnitude (mg) | time | 0.396 | 0.000 | * | 0.445 | 0.000 | * |
| Ankle magnitude (mg) | Sex:time | 0.396 | 0.039 | * | 0.445 | 0.032 | * |
| Wrist magnitude (mg) | time | 0.347 | 0.000 | * | 0.384 | 0.000 | * |
| Wrist magnitude (mg) | Sex:time | 0.347 | 0.432 |  | 0.384 | 0.435 |  |

**Table 11:** Unstandardized LM-estimates for male participants. Values of ankle and wrist ENMO are indicated in mg per day and activities in minutes per day. Stars indicate the significance, one-star p*<* 0.05, two stars p*<* 0.01. Signs of *−* and + indicate the beneficial direction per health marker.

| Health Marker | Ankle ENMO | Wrist ENMO | Active | Household | Sedentary |
| --- | --- | --- | --- | --- | --- |
| Systolic BP (–) | -0.02 (0.894) | -0.06 (0.455) | -0.02 (0.535) | 0.01 (0.741) | -0.0 (0.922) |
| Diastolic BP (–) | 0.03 (0.902) | -0.11 (0.489) | -0.01 (0.841) | -0.05 (0.47) | 0.05 (0.59) |
| Weight (–) | -0.12 (0.538) | -0.18 (0.155) | -0.01 (0.788) | -0.05 (0.337) | 0.05 (0.486) |
| Body Mass Index (–) | -0.19 (0.816) | -0.36 (0.501) | -0.11 (0.52) | -0.36 (0.127) | 0.41 (0.2) |
| Waist circumference (–) | -0.3 (0.144) | -0.29 (0.032)* | -0.04 (0.32) | -0.09 (0.124) | 0.14 (0.087) |
| Waist hip ratio (–) | -53.65 (0.069) | -31.14 (0.107) | -11.58 (0.072) | -10.38 (0.232) | 24.24 (0.038)* |
| Whole body fat (–) | -0.68 (0.009)** | -0.47 (0.005)** | -0.15 (0.01)* | -0.08 (0.294) | 0.27 (0.009)** |
| Trunk fat (–) | -0.69 (0.007)** | -0.49 (0.003)** | -0.16 (0.004)** | -0.08 (0.287) | 0.28 (0.007)** |
| Glycerol (–) | -164.0 (0.041)* | -86.99 (0.099) | -38.73 (0.026)* | -19.64 (0.408) | 75.0 (0.018)* |
| Glycoprotein acetyls (–) | -28.92 (0.011)* | -16.43 (0.029)* | -6.25 (0.012)* | -3.86 (0.261) | 11.4 (0.012)* |
| C-Reactive Protein (–) | 0.52 (0.69) | 0.43 (0.609) | 0.02 (0.929) | 0.51 (0.175) | -0.6 (0.244) |
| Glucose (–) | 0.13 (0.974) | 2.12 (0.379) | -0.02 (0.977) | 0.62 (0.569) | -0.69 (0.64) |
| Insulin (–) | -1.03 (0.016)* | -0.84 (0.002)** | -0.2 (0.04)* | -0.25 (0.05) | 0.45 (0.008)** |
| Framingham Risk Score (–) | -0.97 (0.122) | -0.61 (0.132) | -0.18 (0.197) | -0.13 (0.469) | 0.3 (0.228) |
| Total triglycerides (–) | -9.41 (0.016)* | -5.36 (0.038)* | -1.9 (0.027)* | -2.69 (0.019)* | 4.92 (0.001)** |
| LDL Cholesterol (–) | -5.78 (0.181) | -2.57 (0.364) | -0.87 (0.361) | -0.32 (0.798) | 1.47 (0.394) |
| LDL Diameter (+) | 19.44 (0.326) | -10.92 (0.397) | 0.8 (0.854) | 2.69 (0.644) | -6.26 (0.427) |
| HDL Cholesterol (+) | 9.32 (0.197) | 9.35 (0.044)* | 1.71 (0.279) | 5.28 (0.01)* | -6.8 (0.015)* |
| HDL Diameter (+) | 20.05 (0.015)* | 12.7 (0.018)* | 3.42 (0.06) | 4.71 (0.053) | -8.33 (0.011)* |

**Figure 8:**
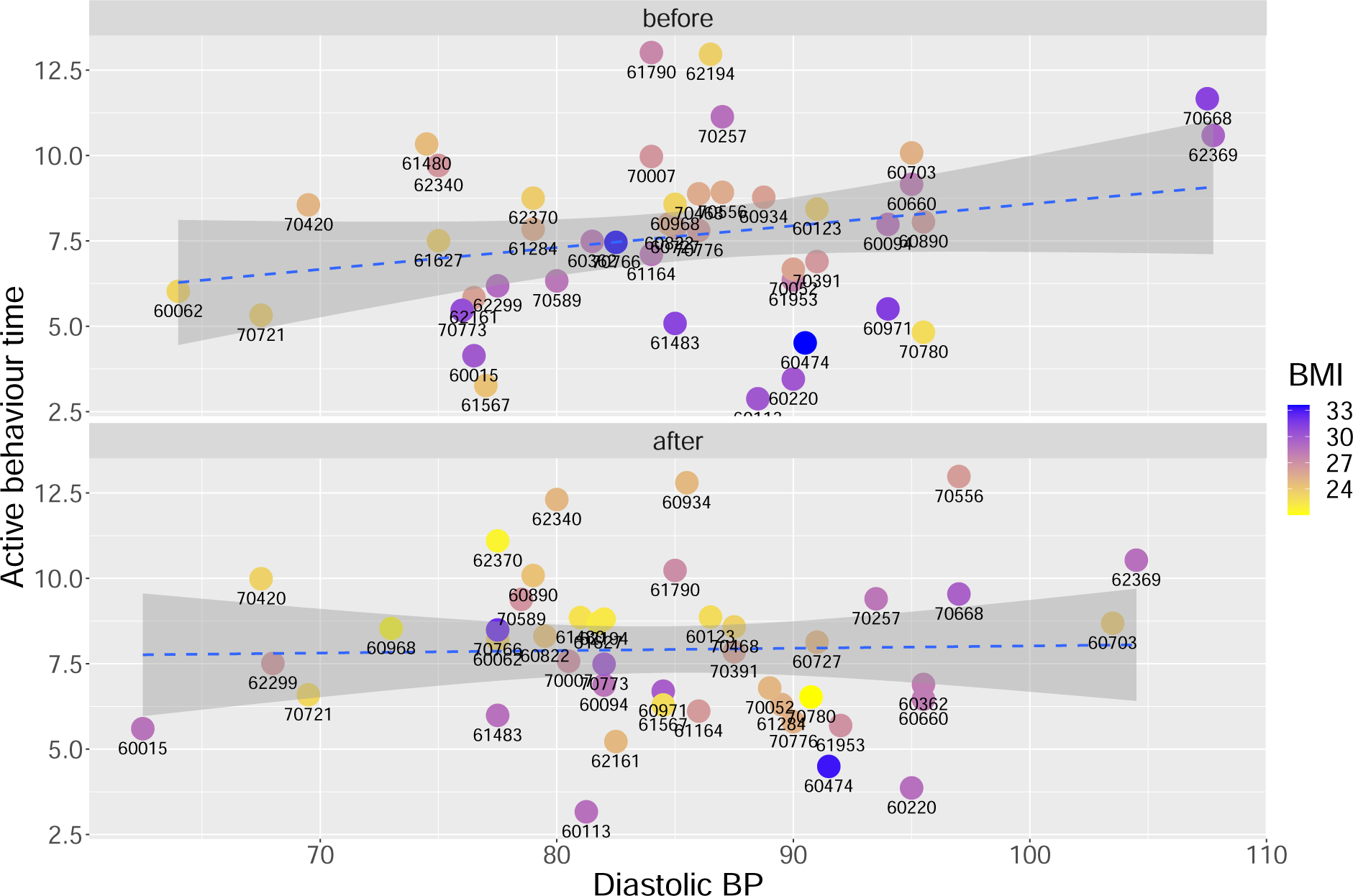
Relationship between blood pressure parameters, BMI, and active behaviour (includes the time spent in walking or cycling) for women at baseline. The numbers next to every point are the participants’ IDs. From Figure 8, the significant association of PB with higher diastolic blood pressure at baseline, can be attributed to our small sample size, where the mean effect size is heavily influenced by two outliers. These outliers consist of two highly active women with very high baseline BMI and elevated blood pressure.

**Table 12:** Unstandardized LM-estimates for female participants. Values of ankle and wrist ENMO are indicated in mg per day and activities in minutes per day. Stars indicate the significance, one-star p*<* 0.05, two stars p*<* 0.01. Signs of *−* and + indicate the beneficial direction per health marker.

| Health Marker | Ankle ENMO | Wrist ENMO | Active | Household | Sedentary |
| --- | --- | --- | --- | --- | --- |
| Systolic BP (−) | 0.18 (0.062) | 0.02 (0.871) | 0.04 (0.086) | 0.03 (0.407) | -0.07 (0.111) |
| Diastolic BP (−) | 0.46 (0.033)* | 0.15 (0.499) | 0.11 (0.046)* | 0.13 (0.123) | -0.24 (0.015)* |
| Weight (−) | -0.12 (0.469) | -0.29 (0.069) | -0.03 (0.406) | -0.08 (0.177) | 0.08 (0.292) |
| Body Mass Index (−) | -0.77 (0.257) | -1.33 (0.041)* | -0.27 (0.103) | -0.42 (0.102) | 0.44 (0.156) |
| Waist circumference (−) | -0.18 (0.323) | -0.28 (0.11) | -0.05 (0.272) | -0.04 (0.533) | 0.06 (0.471) |
| Waist hip ratio (−) | -3.44 (0.857) | -4.73 (0.797) | -1.98 (0.675) | -0.54 (0.943) | 1.0 (0.912) |
| Whole body fat (−) | -0.39 (0.064) | -0.45 (0.031)* | -0.11 (0.033)* | -0.18 (0.02)* | 0.26 (0.006)** |
| Trunk fat (−) | -0.36 (0.094) | -0.39 (0.065) | -0.12 (0.022)* | -0.14 (0.089) | 0.22 (0.026)* |
| Glycerol (−) | -113.0 (0.037)* | -94.45 (0.078) | -38.44 (0.003)** | -29.46 (0.159) | 60.08 (0.016)* |
| Glycoprotein acetyls (−) | -10.78 (0.316) | -4.75 (0.653) | -2.5 (0.344) | -4.93 (0.226) | 6.01 (0.227) |
| C-Reactive Protein (−) | -0.2 (0.845) | 0.6 (0.552) | 0.08 (0.75) | -0.56 (0.142) | 0.4 (0.397) |
| Glucose (−) | -3.72 (0.186) | -4.08 (0.138) | -0.94 (0.179) | -0.92 (0.391) | 0.97 (0.459) |
| Insulin (−) | -0.63 (0.097) | -0.38 (0.32) | -0.17 (0.071) | -0.3 (0.038)* | 0.37 (0.035)* |
| Framingham Risk Score (−) | -0.29 (0.617) | -0.45 (0.424) | -0.06 (0.66) | -0.26 (0.24) | 0.24 (0.359) |
| Total triglycerides (−) | -5.55 (0.131) | -2.7 (0.459) | -1.3 (0.152) | -1.7 (0.226) | 2.52 (0.142) |
| LDL Cholesterol (−) | 0.21 (0.957) | -2.75 (0.461) | -0.51 (0.583) | -1.65 (0.252) | 1.9 (0.28) |
| LDL Diameter (+) | 17.27 (0.31) | 3.14 (0.851) | 5.37 (0.201) | 0.63 (0.924) | -5.04 (0.525) |
| HDL Cholesterol (+) | 9.44 (0.122) | 6.72 (0.266) | 1.35 (0.376) | 2.97 (0.204) | -2.77 (0.331) |
| HDL Diameter (+) | 11.12 (0.143) | 8.73 (0.243) | 2.23 (0.235) | 3.44 (0.235) | -3.77 (0.287) |

**Table 13:**
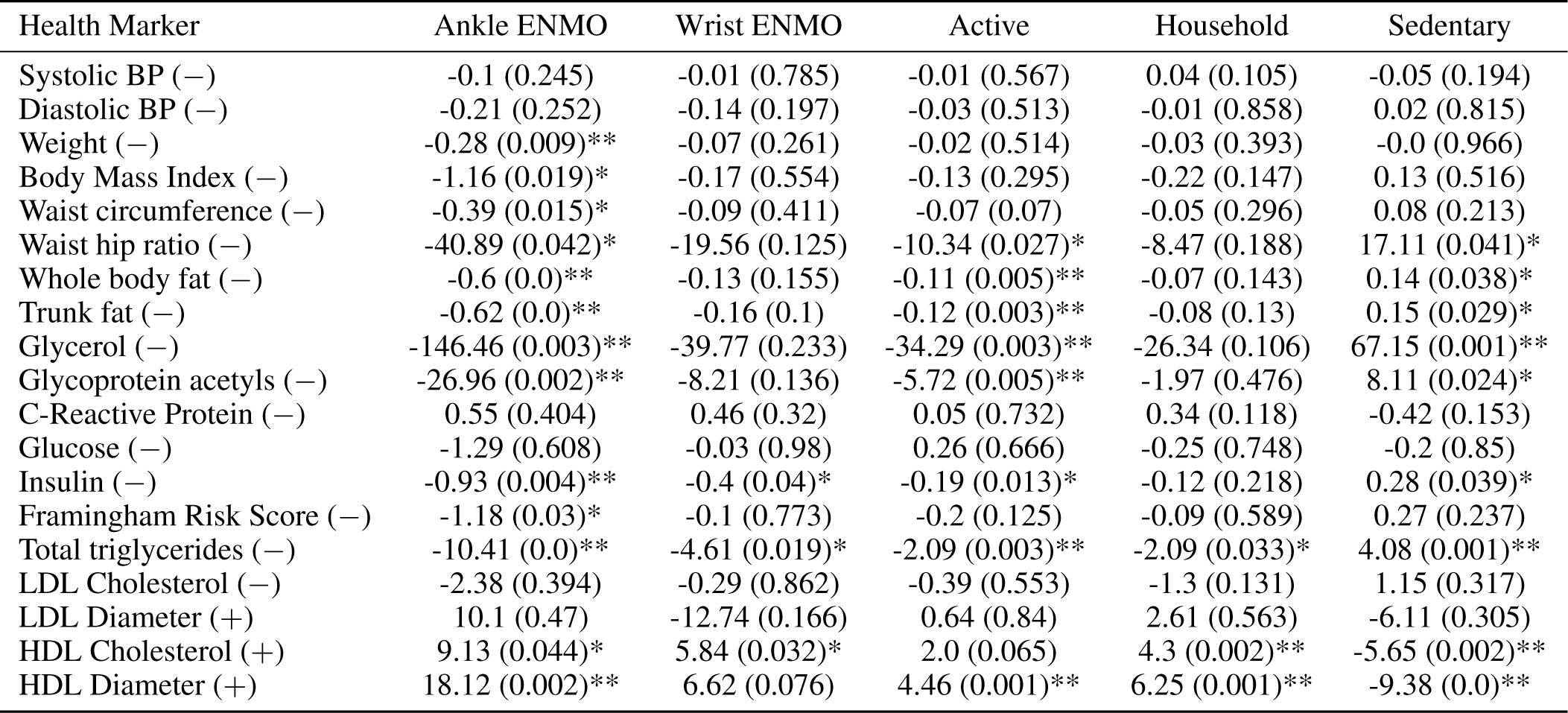
Unstandardized LMM-estimates for male participants. Values of ankle and wrist ENMO are indicated in mg per day and activities in minutes per day. Stars indicate the significance, one-star p*<* 0.05, two stars p*<* 0.01. Signs of *−* and + indicate the beneficial direction per health marker.

| Health Marker | Ankle ENMO | Wrist ENMO | Active | Household | Sedentary |
| --- | --- | --- | --- | --- | --- |
| Systolic BP (−) | -0.1 (0.245) | -0.01 (0.785) | -0.01 (0.567) | 0.04 (0.105) | -0.05 (0.194) |
| Diastolic BP (−) | -0.21 (0.252) | -0.14 (0.197) | -0.03 (0.513) | -0.01 (0.858) | 0.02 (0.815) |
| Weight (−) | -0.28 (0.009)** | -0.07 (0.261) | -0.02 (0.514) | -0.03 (0.393) | -0.0 (0.966) |
| Body Mass Index (−) | -1.16 (0.019)* | -0.17 (0.554) | -0.13 (0.295) | -0.22 (0.147) | 0.13 (0.516) |
| Waist circumference (−) | -0.39 (0.015)* | -0.09 (0.411) | -0.07 (0.07) | -0.05 (0.296) | 0.08 (0.213) |
| Waist hip ratio (−) | -40.89 (0.042)* | -19.56 (0.125) | -10.34 (0.027)* | -8.47 (0.188) | 17.11 (0.041)* |
| Whole body fat (−) | -0.6 (0.0)** | -0.13 (0.155) | -0.11 (0.005)** | -0.07 (0.143) | 0.14 (0.038)* |
| Trunk fat (−) | -0.62 (0.0)** | -0.16 (0.1) | -0.12 (0.003)** | -0.08 (0.13) | 0.15 (0.029)* |
| Glycerol (−) | -146.46 (0.003)** | -39.77 (0.233) | -34.29 (0.003)** | -26.34 (0.106) | 67.15 (0.001)** |
| Glycoprotein acetyls (−) | -26.96 (0.002)** | -8.21 (0.136) | -5.72 (0.005)** | -1.97 (0.476) | 8.11 (0.024)* |
| C-Reactive Protein (−) | 0.55 (0.404) | 0.46 (0.32) | 0.05 (0.732) | 0.34 (0.118) | -0.42 (0.153) |
| Glucose (−) | -1.29 (0.608) | -0.03 (0.98) | 0.26 (0.666) | -0.25 (0.748) | -0.2 (0.85) |
| Insulin (−) | -0.93 (0.004)** | -0.4 (0.04)* | -0.19 (0.013)* | -0.12 (0.218) | 0.28 (0.039)* |
| Framingham Risk Score (−) | -1.18 (0.03)* | -0.1 (0.773) | -0.2 (0.125) | -0.09 (0.589) | 0.27 (0.237) |
| Total triglycerides (−) | -10.41 (0.0)** | -4.61 (0.019)* | -2.09 (0.003)** | -2.09 (0.033)* | 4.08 (0.001)** |
| LDL Cholesterol (−) | -2.38 (0.394) | -0.29 (0.862) | -0.39 (0.553) | -1.3 (0.131) | 1.15 (0.317) |
| LDL Diameter (+) | 10.1 (0.47) | -12.74 (0.166) | 0.64 (0.84) | 2.61 (0.563) | -6.11 (0.305) |
| HDL Cholesterol (+) | 9.13 (0.044)* | 5.84 (0.032)* | 2.0 (0.065) | 4.3 (0.002)** | -5.65 (0.002)** |
| HDL Diameter (+) | 18.12 (0.002)** | 6.62 (0.076) | 4.46 (0.001)** | 6.25 (0.001)** | -9.38 (0.0)** |

**Table 14:** Unstandardized LMM-estimates for female participants. Values of ankle and wrist ENMO are indicated in mg per day and activities in minutes per day. Stars indicate the significance, one-star p*<* 0.05, two stars p*<* 0.01. Signs of *−* and + indicate the beneficial direction per health marker.

| Health Marker | Ankle ENMO | Wrist ENMO | Active | Household | Sedentary |
| --- | --- | --- | --- | --- | --- |
| Systolic BP (–) | 0.04 (0.545) | -0.02 (0.686) | 0.01 (0.348) | 0.01 (0.715) | -0.03 (0.395) |
| Diastolic BP (–) | 0.09 (0.532) | -0.06 (0.593) | 0.03 (0.367) | 0.03 (0.63) | -0.09 (0.268) |
| Weight (–) | -0.12 (0.073) | -0.18 (0.0)** | -0.03 (0.093) | -0.03 (0.371) | 0.04 (0.297) |
| Body Mass Index (–) | -0.67 (0.037)* | -0.88 (0.0)** | -0.2 (0.024)* | -0.15 (0.296) | 0.24 (0.187) |
| Waist circumference (–) | -0.18 (0.16) | -0.32 (0.0)** | -0.05 (0.138) | -0.07 (0.175) | 0.1 (0.152) |
| Waist hip ratio (–) | 6.22 (0.743) | -14.2 (0.313) | 2.63 (0.576) | -2.62 (0.714) | -1.92 (0.835) |
| Whole body fat (–) | -0.31 (0.01)* | -0.3 (0.0)** | -0.08 (0.007)** | -0.06 (0.237) | 0.13 (0.055) |
| Trunk fat (–) | -0.31 (0.007)** | -0.26 (0.001)** | -0.09 (0.004)** | -0.02 (0.668) | 0.08 (0.199) |
| Glycerol (–) | -31.0 (0.486) | -65.24 (0.04)* | -30.91 (0.004)** | -25.89 (0.129) | 57.4 (0.009)** |
| Glycoprotein acetyls (–) | -7.28 (0.309) | -1.87 (0.715) | -0.28 (0.876) | -3.6 (0.208) | 3.65 (0.33) |
| C-Reactive Protein (–) | -0.19 (0.756) | -0.37 (0.415) | -0.04 (0.775) | -0.29 (0.227) | 0.26 (0.413) |
| Glucose (–) | -2.72 (0.195) | -1.11 (0.472) | -0.84 (0.106) | -0.38 (0.639) | 0.72 (0.491) |
| Insulin (–) | -0.33 (0.169) | -0.05 (0.749) | -0.13 (0.042)* | -0.09 (0.365) | 0.19 (0.142) |
| Framingham Risk Score (–) | -0.62 (0.104) | -0.44 (0.106) | -0.03 (0.733) | -0.25 (0.105) | 0.28 (0.172) |
| Total triglycerides (–) | -3.76 (0.109) | -2.15 (0.196) | -0.1 (0.868) | -1.55 (0.104) | 1.67 (0.177) |
| LDL Cholesterol (–) | -2.74 (0.217) | -0.47 (0.768) | -0.21 (0.713) | -0.3 (0.751) | 0.24 (0.837) |
| LDL Diameter (+) | 6.62 (0.573) | 6.12 (0.476) | 2.72 (0.359) | 2.42 (0.603) | -4.79 (0.426) |
| HDL Cholesterol (+) | -0.69 (0.808) | -0.32 (0.863) | 0.32 (0.666) | 1.64 (0.177) | -1.8 (0.259) |
| HDL Diameter (+) | 3.72 (0.324) | 3.44 (0.193) | 0.81 (0.422) | 1.54 (0.345) | -1.61 (0.452) |

**Table 15:** Unstandardized LMM-estimates active behaviour activities and male participants. Values are indicated in minutes per day. Stars indicate the significance, one-star p*<* 0.05, two stars p*<* 0.01. Signs of *−* and + indicate the beneficial direction per health marker.

| Health Marker | Walking | Cycling | Dishwashing | Staking Shelves | Vacuum Cleaning |
| --- | --- | --- | --- | --- | --- |
| Systolic BP (–) | -0.02 (0.128) | 0.01 (0.337) | 0.0 (0.548) | 0.04 (0.102) | 0.01 (0.489) |
| Diastolic BP (–) | -0.04 (0.183) | 0.01 (0.572) | -0.0 (0.779) | 0.02 (0.619) | -0.01 (0.48) |
| Weight (–) | -0.0 (0.889) | -0.01 (0.465) | -0.0 (0.86) | -0.04 (0.102) | 0.0 (0.739) |
| Body Mass Index (–) | -0.01 (0.919) | -0.08 (0.172) | -0.01 (0.803) | -0.25 (0.04)* | -0.01 (0.862) |
| Waist circumference (–) | -0.02 (0.517) | -0.04 (0.047)* | 0.01 (0.555) | -0.07 (0.103) | -0.01 (0.704) |
| Waist hip ratio (–) | -1.85 (0.559) | -6.22 (0.009)** | 0.32 (0.854) | -9.39 (0.065) | -0.9 (0.675) |
| Whole body fat (–) | -0.04 (0.101) | -0.03 (0.079) | -0.01 (0.327) | -0.02 (0.525) | -0.02 (0.176) |
| Trunk fat (–) | -0.05 (0.05) | -0.03 (0.101) | -0.01 (0.353) | -0.02 (0.578) | -0.03 (0.115) |
| Glycerol (–) | -19.4 (0.012)* | -10.75 (0.085) | 3.89 (0.388) | -8.62 (0.509) | -14.77 (0.007)** |
| Glycoprotein acetyls (–) | -3.66 (0.006)** | -1.12 (0.285) | 0.08 (0.918) | -0.32 (0.887) | -1.07 (0.249) |
| C-Reactive Protein (–) | 0.04 (0.719) | 0.01 (0.892) | 0.01 (0.854) | 0.14 (0.422) | 0.15 (0.052) |
| Glucose (–) | -0.15 (0.683) | 0.32 (0.26) | 0.13 (0.517) | -0.54 (0.375) | -0.01 (0.982) |
| Insulin (–) | -0.12 (0.012)* | -0.02 (0.549) | 0.02 (0.414) | -0.09 (0.26) | -0.05 (0.168) |
| Framingham Risk Score (–) | -0.17 (0.042)* | 0.0 (0.982) | -0.04 (0.398) | 0.01 (0.945) | -0.04 (0.55) |
| Total triglycerides (–) | -1.4 (0.003)** | -0.43 (0.249) | -0.16 (0.548) | -0.92 (0.237) | -0.74 (0.027)* |
| LDL Cholesterol (–) | -0.27 (0.526) | -0.06 (0.851) | -0.37 (0.096) | -0.72 (0.293) | -0.16 (0.565) |
| LDL Diameter (+) | 0.26 (0.904) | 0.35 (0.843) | 2.26 (0.071) | 0.65 (0.855) | -0.03 (0.984) |
| HDL Cholesterol (+) | 0.85 (0.219) | 0.66 (0.202) | 0.02 (0.948) | 2.74 (0.014)* | 1.18 (0.009)** |
| HDL Diameter (+) | 1.5 (0.105) | 2.07 (0.003)** | 0.57 (0.25) | 4.02 (0.007)** | 1.32 (0.034)* |

**Table 16:** Unstandardized LMM-estimates active behaviour activities and female participants. Values are indicated in minutes per day. Stars indicate the significance, one-star p*<* 0.05, two stars p*<* 0.01. Signs of *−* and + indicate the beneficial direction per health marker.

| Health Marker | Walking | Cycling | Dishwashing | Staking Shelves | Vacuum Cleaning |
| --- | --- | --- | --- | --- | --- |
| Systolic BP (–) | 0.0 (0.859) | 0.01 (0.257) | -0.0 (0.576) | 0.01 (0.472) | -0.0 (0.962) |
| Diastolic BP (–) | 0.04 (0.146) | -0.01 (0.725) | -0.01 (0.49) | 0.04 (0.36) | -0.0 (0.961) |
| Weight (–) | -0.02 (0.116) | -0.0 (0.63) | 0.0 (0.98) | -0.03 (0.173) | 0.0 (0.622) |
| Body Mass Index (–) | -0.1 (0.072) | -0.05 (0.308) | 0.01 (0.831) | -0.17 (0.126) | 0.01 (0.808) |
| Waist circumference (–) | -0.04 (0.083) | -0.0 (0.896) | 0.0 (0.786) | -0.07 (0.084) | -0.01 (0.737) |
| Waist hip ratio (–) | -1.54 (0.627) | 3.68 (0.161) | 1.58 (0.462) | -3.92 (0.48) | -0.62 (0.81) |
| Whole body fat (–) | -0.03 (0.198) | -0.04 (0.025)* | 0.01 (0.556) | -0.07 (0.063) | 0.0 (0.929) |
| Trunk fat (–) | -0.03 (0.081) | -0.03 (0.047)* | 0.01 (0.58) | -0.03 (0.378) | 0.0 (0.855) |
| Glycerol (–) | -14.86 (0.041)* | -10.13 (0.096) | -1.18 (0.824) | -18.75 (0.157) | -6.69 (0.275) |
| Glycoprotein acetyls (–) | -1.32 (0.267) | 1.11 (0.261) | -1.13 (0.213) | -2.28 (0.302) | -0.14 (0.891) |
| C-Reactive Protein (–) | -0.03 (0.794) | -0.01 (0.908) | 0.0 (0.991) | -0.2 (0.283) | -0.1 (0.267) |
| Glucose (–) | -0.69 (0.048)* | -0.04 (0.882) | 0.35 (0.153) | -0.54 (0.383) | -0.24 (0.411) |
| Insulin (–) | -0.05 (0.251) | -0.05 (0.133) | 0.0 (0.89) | -0.11 (0.15) | 0.01 (0.771) |
| Framingham Risk Score (–) | -0.08 (0.224) | 0.05 (0.346) | 0.0 (0.972) | -0.17 (0.143) | -0.07 (0.192) |
| Total triglycerides (–) | -0.5 (0.206) | 0.38 (0.231) | -0.77 (0.01)* | -0.6 (0.41) | -0.11 (0.737) |
| LDL Cholesterol (–) | -0.03 (0.928) | -0.14 (0.641) | -0.34 (0.25) | 0.33 (0.638) | -0.22 (0.507) |
| LDL Diameter (+) | -0.84 (0.672) | 2.98 (0.066) | 0.54 (0.712) | 1.4 (0.696) | 0.47 (0.774) |
| HDL Cholesterol (+) | -0.09 (0.844) | 0.32 (0.406) | 0.8 (0.046)* | 0.56 (0.549) | 0.31 (0.468) |
| HDL Diameter (+) | 0.12 (0.854) | 0.47 (0.369) | 1.31 (0.014)* | -0.28 (0.822) | 0.55 (0.344) |

## Footnotes

7 www.trialregister.nl

8 GENEActiv Original; Activinsights Ltd, Kimbolton, Cambs, UK, http://www.geneactiv.org/

9 g units; 1 g = 9.81 m/s^2^; 1 mg = 0.00981 m/s^2^

10 The *z*-transform was used: subtracting the mean and dividing by the standard deviation.

11 We compared our results to the activities shown in Table 2 from UKB [13]. For this reason, sitting and standing are merged into one activity

12 1 mg increase of ankle ENMO in men is approx. a 3% increase per day, see Table 3.

13 1 mg increase of wrist ENMO in women is approx. a 3.5% increase per day, see Table 3.

